# A method for lung cancer detection and staging from a drop of blood plasma via Raman spectroscopy of well-based samples (ROWS)

**DOI:** 10.64898/2025.12.17.25341841

**Authors:** Katherine J. I. Ember, Frédérick Dallaire, Éloise D’Amours, Marwa Bounaas, Esmat Zamani, Romane Le Roy-Pépin, Nassim Ksantini, Guillaume Sheehy, Francois Daoust, Juliette Selb, Moishe Liberman, Dominique Trudel, Frédéric Leblond

**Affiliations:** Polytechnique Montréal, Montréal, Canada; Centre de recherche du Centre hospitalier de l’Université de Montréal, Montréal, Canada; Université de Montréal, Montréal, Canada; Reveal Life Science, Montréal, Canada; Centre hospitalier de l’Université de Montréal, Montréal, Canada; Institut du cancer de Montréal, Montreal, Canada; Department of Molecular Biology, Medical Biochemistry and Pathology, Université Laval, Quebec City, Canada; Quebec Heart & Lung Institute Research Center, Quebec City, Canada

## Abstract

We present a new method for lung pathology detection in blood plasma, including lung cancer staging. Raman spectroscopy uses inelastically scattered laser light to obtain molecular information in a reagent-free manner. Obtaining Raman spectral data from liquid samples has long proven challenging, but we have developed a novel tool for obtaining spectra from 60 μl liquid samples within two minutes: Raman of Well-based Samples (ROWS). With a low-cost ROWS device, we analyzed 372 blood plasma samples from a national biobank, including controls (n=92), patients with stage I-II lung cancer (n=99), stage III-IV cancer (n=46), benign tumours (n=36) and other lung conditions (n=99). Machine learning models were built to assess lung cancer stage and lung pathology presence. ROWS achieves up to 94% sensitivity, 90% specificity and 93% accuracy depending on classification. ROWS proves a robust method for rapid, low-cost, user-friendly, point-of-care lung pathology analysis in small quantities of blood plasma.

**One Sentence Summary:** A new way of detecting lung cancer in small volumes of liquid blood plasma was developed using laser-based Raman spectroscopy and metallic wells.

## INTRODUCTION

Lung cancer is the leading cause of cancer mortality globally, accounting for almost as many deaths per year as the second, third and fourth deadliest cancers combined: colorectal, liver and breast cancer (*1*). This is partly due to inefficient lung cancer detection practices leading to late-stage diagnosis (*2*). Portable, rapid, inexpensive, non-ionizing techniques could greatly benefit lung cancer detection, and yet no such tools exist in the clinic today.

Lung cancer is classified as either non-small cell lung cancer (NSCLC, 85% of lung cancer cases) or small cell lung cancer (SCLC, 15% of cases) (*3*). Five-year recurrence free survival rates for NSCLC generally depend on the cancer stage at diagnosis: Stage I: 86%, Stage II: 50%, Stage III: 34% (*4*). Lung cancer screening programs have been shown to improve survival rates by reducing the stage at which lung cancer is detected, with Stage I lung cancer making up approximately 65% of detections compared to 14% in non-screening detection, (*5, 6*). Identifying cancer stage also allows clinicians to select the correct treatment plan and communicate prognosis with the patient.

The American Cancer Society recommends annual low dose computed tomography (LDCT) lung cancer screening for people between 50 and 80 years old who have at least a 20 pack-year history of smoking (*7*). However, only 5.8% of the 14.2 million eligible Americans partake in screening (*9*) and this is partly due to lack of access: 61% of US counties with high adult smoking rates are over one hour’s drive-time from a screening center of excellence (*10*). The two gold-standard methods for lung cancer screening: LDCT and chest radiography, are limited in widespread availability. Machines are often expensive, not portable, use ionizing radiation in the form of X-rays, and require specialists such as radiographers for operation and interpretation. A portable, user-friendly, minimally invasive, low-cost, point-of-care test for lung cancer would facilitate screening uptake in remote or low-income locations, potentially reducing mortality. Furthermore, such tests could allow lung cancer staging and permit monitoring patients for lung cancer recurrence. Blood products are a favorite for minimally invasive testing due to high analyte concentrations and low chance of contamination compared to other biofluids. Effective blood-based tests would reduce the number of diagnostic imaging exams, shorten the uncertainty period with indeterminate cases, and reduce patient exposure to potentially harmful ionizing radiation. Such tests could also be integrated into standard medical centers, reducing patient travel time to specialized LDCT centers. Furthermore, some blood plasma biomarker-based tests have been shown to detect cancer 29 months prior to diagnosis using LDCT (*18*). This would allow earlier treatment and potentially better prognosis.

Another use-case for biofluid screening is in the discrimination between benign and malignant lung lesions. This clinical problem leads to the high false positive rates of one-off LDCT and chest radiography imaging: 96.4% and 94.5% respectively (*8*). To address this, the American College of Radiologists have created a standardized screening method: the Lung Computed Tomography Screening Reporting and Data System (Lung-RADS) (*11*). Lung-RADS introduces more frequent LDCT (3-6 months) screening after initial lesion detection, allowing clinicians to assess malignancy based on lesion features and size evolution.

Despite this advance, a recent microsimulation study reported that integration of a non-invasive biochemical screening test costing less than $250 per measurement with a sensitivity of 60% and specificity of 90% for malignant lesions would be cost-effective (*13*). The test was assumed to take place one month after an LDCT exam for those with a Lung-RADS score of 3 or 4A. Candidates for such a molecular test include detection of tumor protein biomarkers such as carbohydrate antigen 19-9 (CA19-9) (*14*). However, these are not specific to lung cancer. Other candidates include techniques based on circulating tumor cells (CTCs),(*15*) CtDNA (DNA shed by tumors of CTCs into the bloodstream), miRNA, tumor derived exosomes and tumor educated platelets (TEPs) (*16*). However, all of these lack standardization and technologies to obtain the target material, (*14*) and CTC equipment costs 600-800K USD (*17*).

A point-of-care device would ideally incorporate the use of small quantities of blood compatible with fingerpricks (less than 100 μl), rapid analysis time (less than 20 min), low cost, and a portable system. Furthermore, for such a test to be used in resource-poor settings, the test would minimize use of reagents or contrast agents as these often require specialized storage conditions and mixing steps by trained personnel.

One possible solution for reagent-free, point-of-care, blood-based analysis is Raman spectroscopy. The technique uses lasers to detect molecular changes, including those associated with diseases.(*19–21*) In Raman spectroscopy, laser light is shone at a sample, and inelastically scattered light results from the interaction of light with molecules in the sample. The Raman scattered light can be detected using a spectrometer. This light contains information about the concentration and identity of molecules in a sample and can be used as a “molecular barcode”. The Raman spectrum of a molecule consists of a series of peaks where the height of the peaks is related to the concentration of the molecule and the position of the peaks on the x-axis is related to the identity of a molecule.(*22*) Raman spectroscopy can be used to gain information about molecular changes in biofluids associated with diseases. This clinically interpretable biomolecular signature may then be used to build biochemically-informed machine learning models to detect disease. Raman spectroscopy can be used to create diagnostic tests without the use of costly, specialised reagents.

A low-cost, non-invasive and rapid lung cancer analysis device using small quantities of liquid blood plasma with minimal level of pre-treatment would be invaluable for the early detection, staging, and treatment of the disease. We developed a method for obtaining high-quality Raman spectra from small quantities of blood plasma compatible with fingerpricks through a custom designed aluminum well. The technique will hereafter be referred to as ROWS (Raman of well-based samples). Here, we present the first proof-of-concept of a ROWS-based device, in this case applied to detection and staging of lung cancer in liquid blood plasma.

## RESULTS

### Raman spectra from liquid blood plasma samples yield biomolecular peaks associated with proteins, lipids and carotenoids

To assess whether the ROWS methodology could be implemented as a lung cancer staging tool, and to screen for other lung pathologies, Raman spectra were obtained from 372 blood plasma samples. This was achieved in a manner compatible with point-of-care biofluid analysis (**Figure 1, Methods**). Blood plasma samples were defrosted, vortexed for mixing, and 60 μl of sample was pipetted into a custom aluminum well. Spectra were obtained from liquid form samples using a user-friendly, single-point Raman spectrometer with no additional requirements for sample positioning other than insertion of the cassette into the device. The time from plasma entering the sample cassette to spectral acquisition was less than 3 minutes for all samples, including sample vortexing (**Methods**).

**Figure 1.**
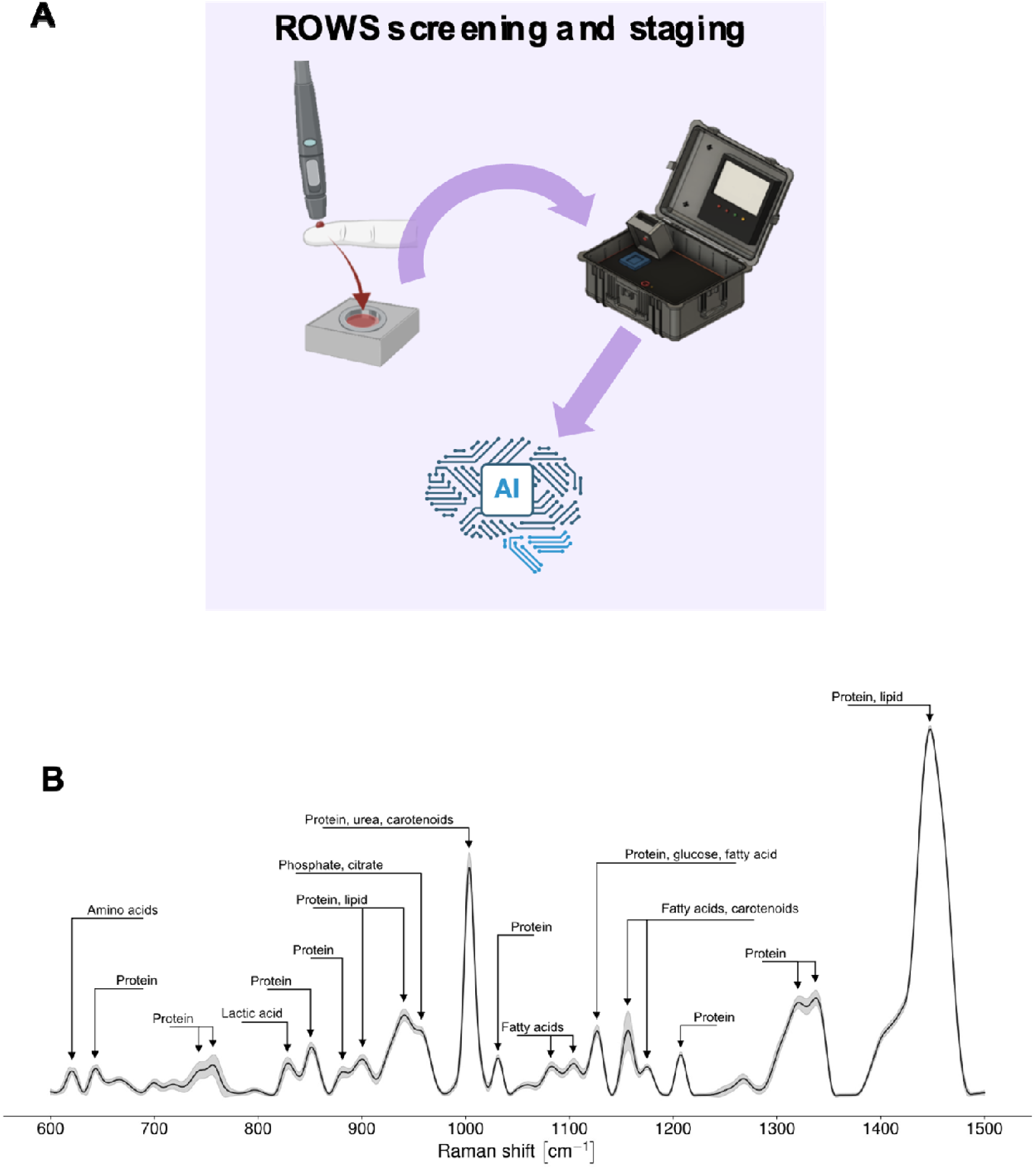
Raman of well-based samples (ROWS) for lung pathology. **(A) Schematic diagram of the ROWS plasma screening and staging tool.** A fingerprick or venous blood extraction is used for sample collection prior to blood processing for plasma. Plasma is inserted into a ROWS samples cassette and then into a portable device. This purple box represents the purple boxes in Figures 2–6. **(B) Mean Raman spectrum of blood plasma from a biobank of lung cancer, controls, and other lung pathologies (n = 372).** Standard deviation is shown by pale grey shading. Tentative biomolecular assignments have been made based on literature.

Blood plasma samples were taken from controls (n=94) and patients with Stage I-II NSCLC (n=99), Stage III-IV NSCLC (n=47), benign lesions (n=36) and other lung conditions (n=99). Non-tumoral lung pathologies included asthma, chronic obstructive pulmonary disease (COPD), interstitial lung disease and pulmonary arterial hypertension (PAH).

ROWS Raman spectra were obtained from each blood plasma sample, with individual peaks associated with multiple biomolecules. The difference between the wavelength of the Raman scattered light from a molecular bond and the wavelength of the incident laser light determines the position of the Raman peak on the x-axis. This value is given in wavenumbers (cm^-1^, inversely proportional to wavelength). The peak height (peak intensity) on the y-axis is associated with the strength of Raman scattering, which is in turn associated with the concentration of the responsible molecules in the illuminated sample.

Changes in Raman spectra between different pathologies and smoking statuses were evaluated. The intensity of each Raman peak was plotted in box-and-whisker plots for twenty pairwise comparisons of patient classes (e.g., control spectra vs. Stage I–II cancer spectra) (**Figures 2E-6E, Supplementary Figures 3C-13C**). Each Raman peak either increased, decreased, or did not change in a statistically significant manner when comparing the first patient class to the second patient class. For each of the twenty comparisons, the response of nineteen Raman peaks were recorded. These responses are shown in **Supplementary Figure 1**. The peak at 1156 cm^-1^ decreased in eight out of twelve comparisons where patients without malignant cancer were the first patient class and patients with malignant cancer constituted the second patient class. It did not change significantly for the remaining four out of twelve non-cancer/cancer comparisons. Meanwhile the peak at 1337 cm^-1^ increased in ten out of twelve comparisons comparisons with non-cancer as the first patient class and malignant cancer as the second patient class. It did not change significantly for the remaining two out of twelve non-cancer/cancer comparisons.

**Figure 2:**
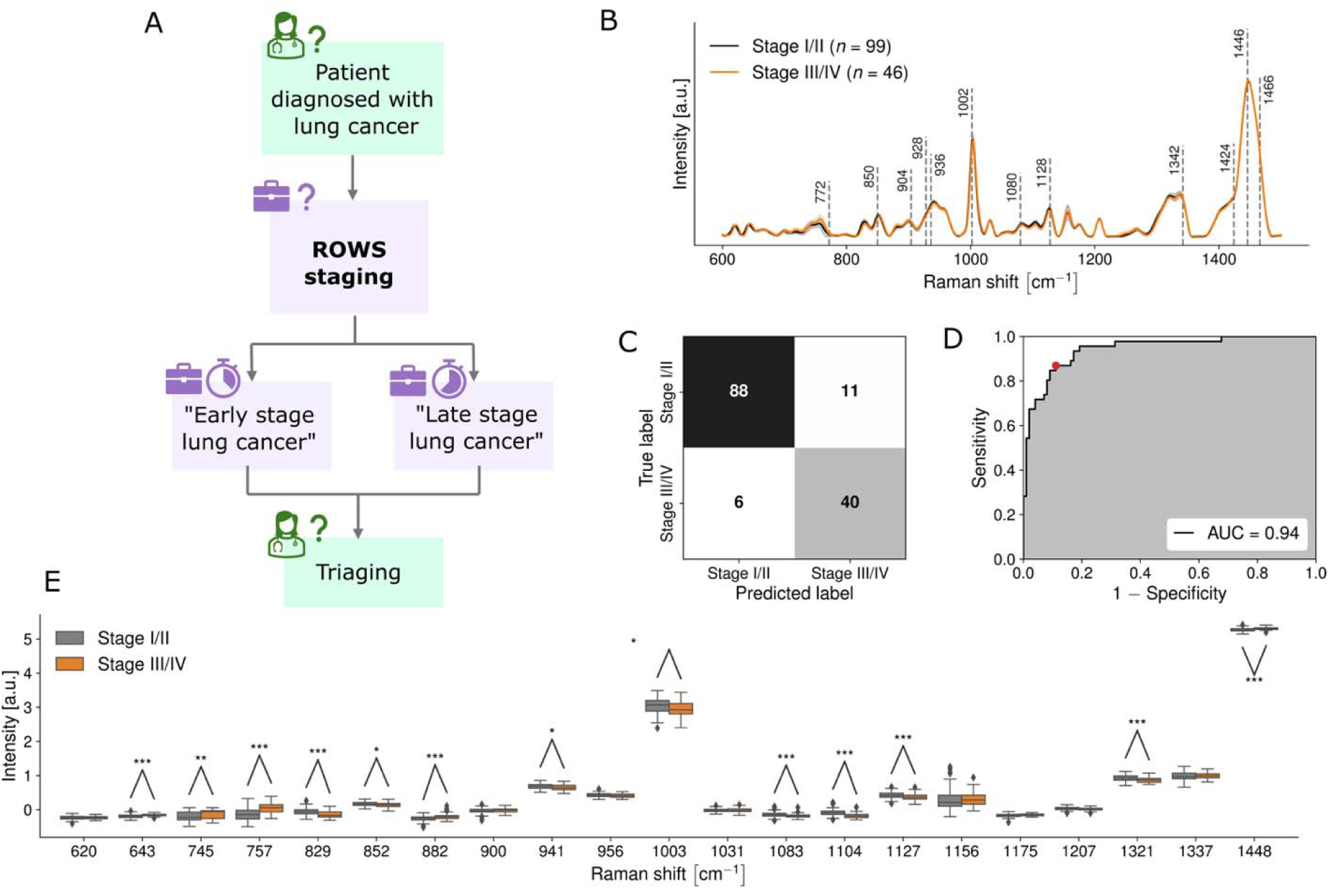
Towards point-of-care lung cancer staging (Model 1): Raman spectroscopy detection of Stage I-II vs. Stage III-IV lung cancers assisted by Raman of well-based samples. (A) Workflows of how a point-of-care ROWS device could be integrated into the clinical workflow. Green boxes indicate boxes where a highly trained clinician is required. Purple boxes indicate the screening and staging device outlined in this manuscript. (B) Raman spectra of liquid blood plasma from patients with Stage I-II (black line) and Stage III-IV lung cancer (orange). Black dashed lines show peaks used in the machine learning models. (B) Confusion matrix showing efficacy of Raman spectral prediction of Stage I-II and Stage III-IV lung cancer in liquid blood plasma. (C) Receiver operating characteristic (ROC) curves for machine learning models discriminating between liquid blood plasma spectral fingerprints from the two groups. (D) Box-and-whisker plot showing the relative intensities of different Raman bands between liquid blood plasma from patients with Stage I-II (grey) and Stage III-IV lung cancer (orange). * represents a p-value of ≤0.05. ** represents a p-value of ≤ 0.01. *** represents a p-value of ≤ 0.001.

To confidently identify the biomolecules responsible for each Raman peak (**Figure 1B**), multiple forms of information were combined. Firstly, literature values of Raman spectra from pure biomolecules were assessed and compared to the plasma Raman spectra (*23–31*). Secondly, correlation values were produced between different pairs of Raman peaks to identify which Raman peaks may belong to the same molecular family. Correlation values were calculated by comparing the directional responses of Raman peaks - whether they increased, decreased, or were unchanged - across multiple predefined patient-class comparisons (**Supplementary Figure 1)**. Peak pairs exhibiting similar response patterns across patient class comparisons were assigned high correlation values (**Supplementary Figure 2**). For example, the 1156 and 1337 cm^-1^ peak pair demonstrated an extremely weak correlation value (-10) as they frequently demonstrated opposing responses to patient pathology and smoking status. This suggests the peaks belong to different biomolecular families. Meanwhile, the 1321 and 1337 cm^-1^ peak pair demonstrated an extremely strong correlation value (10), suggesting that both peaks are associated with the same biomolecular family. Indeed, literature shows that both 1321 and 1337 cm^-1^ are strongly linked to bonds within proteins.

Based on the plasma Raman spectra acquired using the ROWS technique, machine learning models were built for classification of samples (**Table 1 and Supplementary Table 1, Methods**). Models were built for lung cancer staging, lung cancer detection in patients with pulmonary symptoms such as coughing, lung cancer sensing in the general population, detection of non-tumoral lung conditions, and smoking status assessment. “Stage I-IV” samples included those with Stage I-IV lung cancer (n=145). “Healthy controls” were samples without lung pathologies, although they may have had other pathologies not related to the lung such as allergic rhinitis (n=92). “Benign” samples were those with benign lesions in their lungs i.e. lesions that were not malignant cancer (n=36), such as granulomas and hamartomas. “Non-tumoral” pathologies included patients with lung conditions such as asthma, COPD and PAH, but did not include benign lesion or malignant cancer samples (n=99). “Non-cancer” samples included “benign”, “non-tumoral” pathologies and “healthy control” samples (n=227). Machine learning models were built using support vector machines (SVM). To assess machine learning model performance, area under curve of receiver operating characteristic (ROC) curves were calculated. The closer the AUC is to 1, the closer the model is to perfect classification, with 0.5 reflecting random classification. AUCs ranged between 0.56 (ex-smokers with early-stage lung cancer compared to smokers with early-stage lung cancer), and 0.96 (control compared to non-tumoral patients). Accuracies, sensitivities, and specificities were calculated for each model (**Table 2 and Supplementary 2**) for specific points on the AUC curve, determined by clinical need.

**Table 1:** Machine learning models developed using Raman spectra of blood plasma from a biobank acquired from lung cancer patients, healthy controls, and patients with other lung pathologies.

|  | Negative group size | Negative group description | Positive group size | Positive group description |
| --- | --- | --- | --- | --- |
| **Model 1** | 99 | Stage I-II | 46 | Stage III-IV |
| **Model 2** | 99 | Non-tumoral | 145 | Stage I-IV |
| **Model 3** | 92 | Healthy | 145 | Stage I-IV |
| **Model 4** | 92 | Healthy | 99 | Stage I-II |
| **Model 5** | 92 | Healthy | 46 | Stage III-IV |
| **Model 6** | 128 | Healthy/benign | 145 | Stage I-IV |
| **Model 7** | 227 | Non-cancer | 99 | Stage I-II |
| **Model 8** | 227 | Non-cancer | 46 | Stage III-IV |
| **Model 9** | 227 | Non-cancer | 145 | Stage I-IV |
| **Model 10** | 92 | Healthy | 280 | Non-healthy |
| **Model 11** | 92 | Healthy | 99 | Non-tumoral |
| **Model 12** | 36 | Benign | 145 | Stage I-IV |
| **Model 13** | 36 | Benign | 99 | Stage I-II |
| **Model 14** | 36 | Benign | 46 | Stage III-IV |
| **Model 15** | 99 | Non-tumoral | 36 | Benign |
| **Model 16** | 92 | Healthy | 36 | Benign |

**Table 2:**
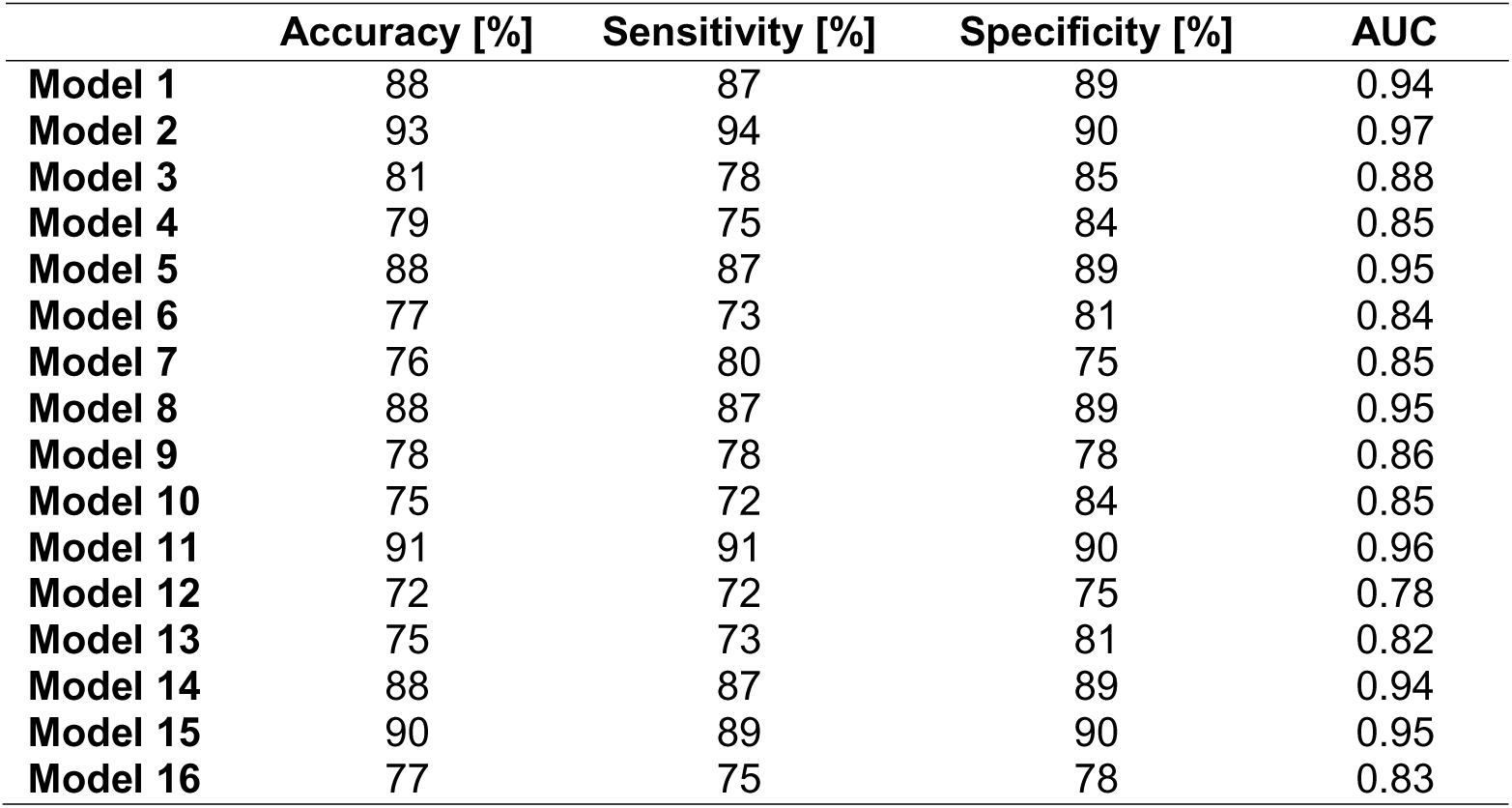
Performance metrics of the machine learning models developed using Raman spectra of blood plasma from a biobank acquired from lung cancer patients, controls, and patients with other lung pathologies.

### ROWS can discriminate between blood plasma samples from early-stage and late-stage cancer

A ROWS-based cancer staging tool could be used to triage patients for urgent care. To establish whether ROWS can be used for cancer staging, a machine learning model was used to classify blood plasma from patients with Stages I-II lung cancer and those with Stages III-IV lung cancer **(Figure 2, Model 1).** This was achieved with 0.94 AUC, 88% accuracy, 87% sensitivity and 89% specificity **(Table 3)**. Models could further be optimized for higher sensitivity, to ensure that all urgent cases are prioritized. Features used in the machine learning model to discriminate between samples included 643, 745, 757, 882 and 1448 cm^-1^ (principally associated with protein) which were higher in later stage cancers relative to earlier stage, and 829, 852, 941, 1003, 1083, 1104, 1127, and 1321 cm^-1^ (carotenoids, fatty acids, certain protein bands) which were lower in later stage cancers. There are no published studies assessing total protein concentration in stage I-II lung cancers compare to stage III-IV lung cancers. However, studies have shown that total serum cholesterol inversely correlated with lung adenocarcinoma survival,(*32*) and the ratio of non-HDL/HDL (high density lipoproteins) negatively correlated with survival.(*33*) Furthermore, multiple studies have shown that serum carotenoid levels are inversely correlated with cancer-related death.(*34, 35*)

**Table 3.** Clinical and pathological information about the cohort of samples under investigation. The number in brackets indicates the percentage of total samples in that group with a particular characteristic. Sample matching was attempted to ensure lack of bias in data.

|  | **Control** | **Non-tumoral** | **Benign** | **Stage I/II** | **Stage III/IV** |
| --- | --- | --- | --- | --- | --- |
| Age, average (std. dev.) | 61 (5) | 62 (5) | 62 (6) | 64 (5) | 66 (5) |
| Sex |  |  |  |  |  |
| Male | 43 (47) | 42 (43) | 18 (50) | 48 (49) | 21 (46) |
| Female | 49 (53) | 53 (54) | 18 (50) | 47 (48) | 24 (52) |
| N/A | — | 3 (3) | — | 2 (2) | 1 (2) |
| Histology, *n* (%) |  |  |  |  |  |
| Squamous cell carcinoma | — | — | — | 26 (27) | 15 (33) |
| Adenocarcinoma | — | — | — | 62 (64) | 26 (57) |
| Neuroendocrine tumor | — | — | — | 6 (6) | 3 (7) |
| Sarcomatoid carcinoma | — | — | — | — | 1 (2) |
| Other | — | — | — | — | 1 (1) |
| Tabagism, *n* (%) |  |  |  |  |  |
| Non-smoker | 41 (45) | 65 (66) | 6 (17) | 8 (8) | — |
| Smoker | 3 (3) | 27 (28) | 4 (11) | 47 (48) | 8 (17) |
| Ex-smoker | 47 (52) | 3 (3) | 25 (69) | 40 (41) | 37 (80) |
| Passive smoker | — | — | 1 (3) | — | — |
| N/A | — | 3 (3) | — | 2 (2) | 1 (2) |
| Ethnicity, *n* (%) |  |  |  |  |  |
| Caucasian | 91 (100) | 95 (97) | 36 (100) | 94 (97) | 45 (98) |
| Indigenous | — | — | — | 1 (1) | — |
| N/A | — | 3 (3) | — | 2 (2) | 1 (2) |

The high sensitivity and specificity of the lung cancer staging model suggested that ROWS could be used as a point-of-care triaging tool after lung cancer diagnosis using the lung-RADS workflow.

### ROWS can discriminate between blood plasma samples from non-tumoral lung pathology patients and lung cancer patients

Non-tumoral lung pathologies may have symptoms similar to lung cancer, for example chronic coughing associated with asthma or chronic obstructive pulmonary disease (COPD). To evaluate whether ROWS could potentially be used to screen for lung cancer amongst other lung pathologies, a machine learning model was built to classify Raman spectra from samples from patients with non-tumoral lung pathologies and those with any lung cancer stage (**Figure 3, Model 2**). Benign lung lesions do not usually cause symptoms, so these were not included. An AUC of 0.97 was achieved, with an accuracy of 93%, a sensitivity of 94% and a specificity of 90% (**Table 3**). Peaks associated with proteins (643, 745, 757, 829, 1321, 1337 and 1448 cm^-1^) increased in cancer samples compared to non-tumoral lung pathologies, whilst peaks from lipids (1083 and 1104 cm^-1^) and carotenoids at 1156 cm^-1^ decreased. Additionally, peaks from phosphate (956 cm^-1^), phenylalanine, protein, and urea (1003 cm^-1^), and a peak due to both proteins and lipids (941 cm^-1^) decreased. NMR spectroscopy-based metabolomics has identified that there is a reduction in lipid levels from blood serum in patients with lung cancer compared to those with COPD, consistent with our results.(*36*) The high specificity and sensitivity of this machine learning model illustrates that ROWS detects the biomolecular signature of lung cancer in blood plasma amongst other lung pathologies independently of the presence of symptoms such as coughing.

**Figure 3:**
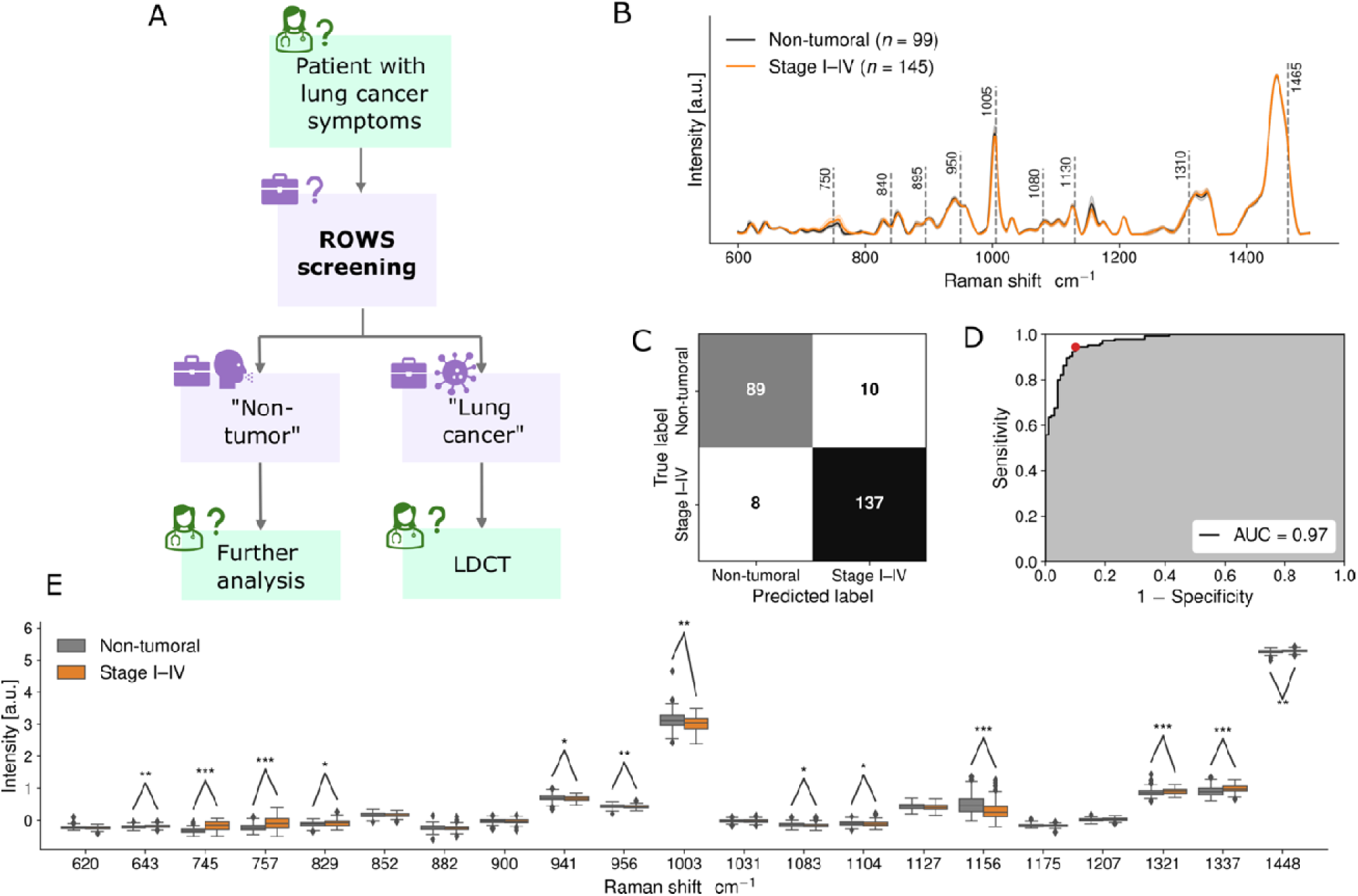
Towards point-of-care lung cancer screening amongst patients with symptoms of coughing (Model 2): Raman spectroscopy detection of non-tumoral vs. Stage I-IV lung cancers assisted by Raman of well-based samples. (A) Workflows of how a point-of-care ROWS device could be integrated into the clinical workflow. Green boxes indicate boxes where a highly trained clinician is required. Purple boxes indicate the screening and staging device outlined in this manuscript. (B) Raman spectra of liquid blood plasma from non-tumoral lung pathologies (black line) and Stage I-IV lung cancer (orange). Black dashed lines show peaks used in the machine learning models. (B) Confusion matrix showing efficacy of Raman spectral prediction of non-tumoral lung pathologies and Stage I-IV lung cancer in liquid blood plasma. (C) Receiver operating characteristic (ROC) curves for machine learning models discriminating between liquid blood plasma spectral fingerprints from the two groups. (D) Box-and-whisker plot showing the relative intensities of different Raman bands between liquid blood plasma from non-tumoral lung pathologies (grey) and those with Stage I-IV lung cancer (orange). * represent a p-value of ≤0.05. ** represents a p-value of ≤ 0.01. *** represents a p-value of ≤ 0.001.

### ROWS can discriminate between blood plasma samples from control volunteers and lung cancer patients

To assess whether ROWS could potentially be used to screen for lung cancer in the general population or to monitor recurrence after treatment, machine learning models were built to discriminate between ROWS measurements from lung cancer patients and controls. Controls did not have lung lesions or lung tumors, or other diagnosed lung conditions. **Model 3,** designed to discriminate between lung cancer regardless of stage and controls did so with an AUC of 0.88, 81% accuracy, 78% sensitivity and 85% specificity **(Figure 4)**. Peaks associated with proteins (620, 643, 745, 757, 829, 852, 882, 900, 1031, 1321, 1337 and 1448 cm^-1^) and lipids (1083 cm^-1^) increased in cancer samples whilst the peak due to carotenoids at 1156 cm^-1^ decreased (**Figure 4D).** There are no published studies assessing total protein concentration in plasma or serum of people with lung cancer. However, research has shown that there are higher total protein concentrations in serum in women with breast cancer compared to healthy controls.(*37*) Studies using high-performance liquid chromatography have shown that higher plasma carotenoids are associated with lower cancer risk for breast cancer.(*38, 39*)

**Figure 4:**
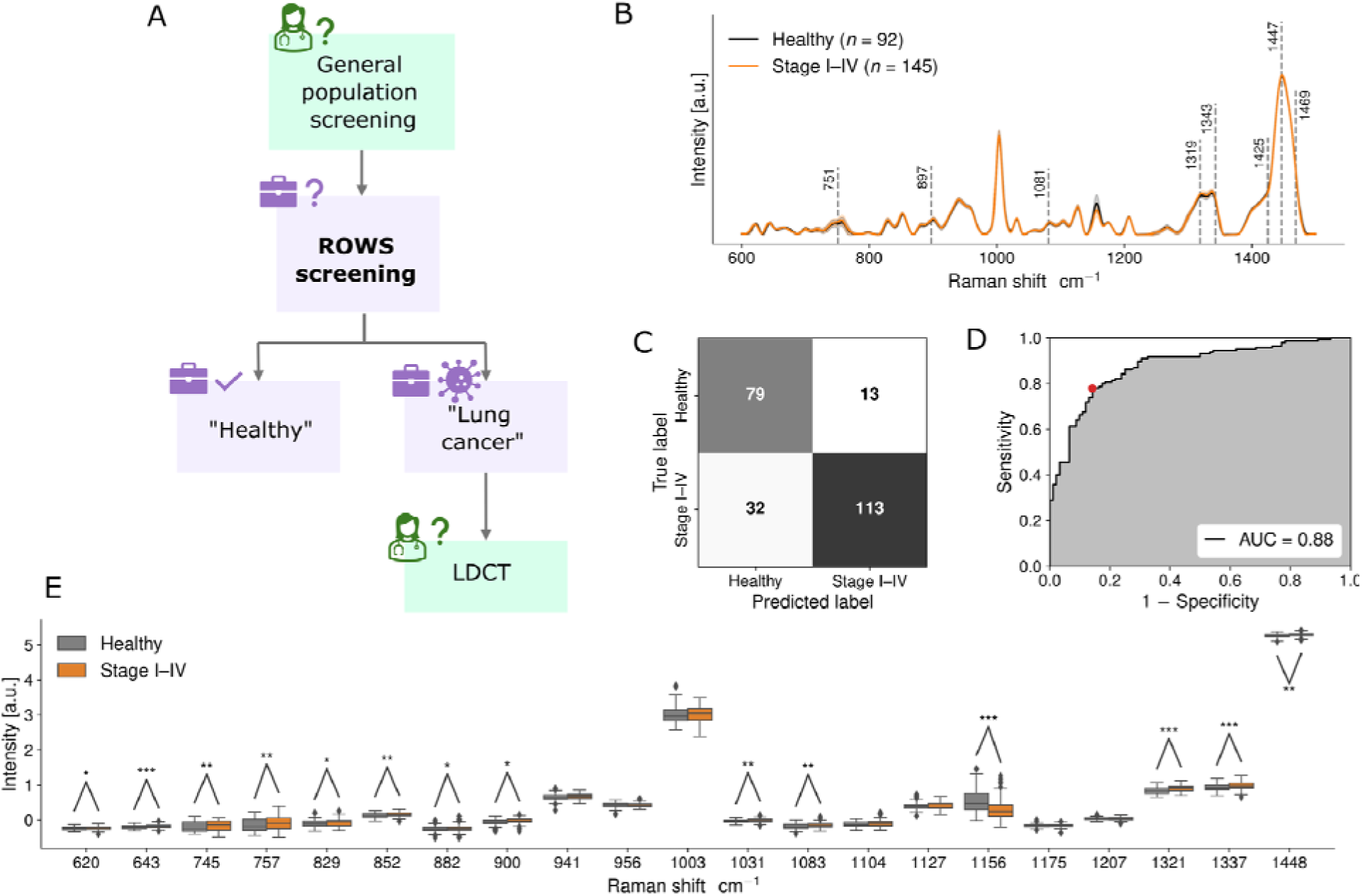
Towards point-of-care lung cancer screening amongst controls assisted by Raman of well-based samples (Model 3): Raman spectroscopy detection of control vs. Stage I-IV lung cancers. (A) Workflows of how a point-of-care ROWS device could be integrated into the clinical workflow. Green boxes indicate boxes where a highly trained clinician is required. Purple boxes indicate the screening and staging device outlined in this manuscript. (B) Raman spectra of liquid blood plasma from healthy controls (black line) and Stage I-IV lung cancer (orange). Black dashed lines show peaks used in the machine learning models. (B) Confusion matrix showing efficacy of Raman spectral prediction of controls and Stage I-IV lung cancer in liquid blood plasma. (C) Receiver operating characteristic (ROC) curves for machine learning models discriminating between liquid blood plasma spectral fingerprints from the two groups. (D) Box-and-whisker plot showing the relative intensities of different Raman bands between liquid blood plasma from control patients (grey) and those with Stage I-IV lung cancer (orange). * represents a p-value of ≤0.05. ** represents a p-value of ≤ 0.01. *** represents a p-value of ≤ 0.001.

**Model 4** classified controls and early stage (I-II) cancers with an AUC of 0.85, accuracy of 79%, a sensitivity of 75% and a specificity of 84% **(Supplementary Figure 3)**. **Model 5** classified healthy controls and late stage (III-IV) cancers with a higher AUC of 0.95, accuracy of 88%, a sensitivity of 87% and a specificity 89% **(Supplementary Figure 4)**. As for the case of benign tumours described above, **Model 5** likely outperformed **Models 6 and 8** as later stage cancer is often associated with more substantial biomolecular changes compared to early-stage cancers.

To assess the efficacy of ROWS when benign lesions were present in the control population, a machine learning model was produced including both controls and those with benign lesions in the negative group, and Stage I-IV lung cancers in the positive group (**Model 6**). This classified with an AUC of 0.84, accuracy of 77%, a sensitivity of 73% and a specificity of 81% **(Supplementary Figure 5)**. To assess the accuracy when benign lesions and other lung pathologies were present at high proportions, machine learning models were built (**Models 7-9**) where all non-cancer samples were used as the negative group and cancer samples were used a the positive group. These ranged from accuracies of 76-88% depending on the stage of cancer **(Table 3, Supplementary Figures 6-8)**.

### ROWS can discriminate between blood plasma samples from healthy controls and lung pathology patients

To investigate whether a patient could be tested for abnormal lung pathology using ROWS, a machine learning model was built to discriminate between controls with no lung pathology and patients with lung pathologies (including lung cancer, benign tumors, asthma, and chronic obstructive pulmonary disease (COPD)). This machine learning model achieved an AUC of 0.85, accuracy of 75%, sensitivity of 72% and specificity of 84% (**Figure 5**, **Table 2, Model 10**). Lung pathologies were associated with an increase in peaks due to proteins (620, 643, 852, 882, 900, 1031, 1321 and 1448 cm^-1^) and lipids (1083, 1104 and 1127 cm^-1^), and a reduction in peaks due to carotenoids (1156 cm^-1^). There was also an increase in the peak at 1003 cm^-1^ due to phenylalanine, protein and urea. Combined with symptoms, ROWS may enhance the diagnosis of lung pathologies. However, further research must be conducted, to assess the sensitivity and specificity of the ROWS device in combination with symptoms such as coughing and difficulty breathing.

**Figure 5:**
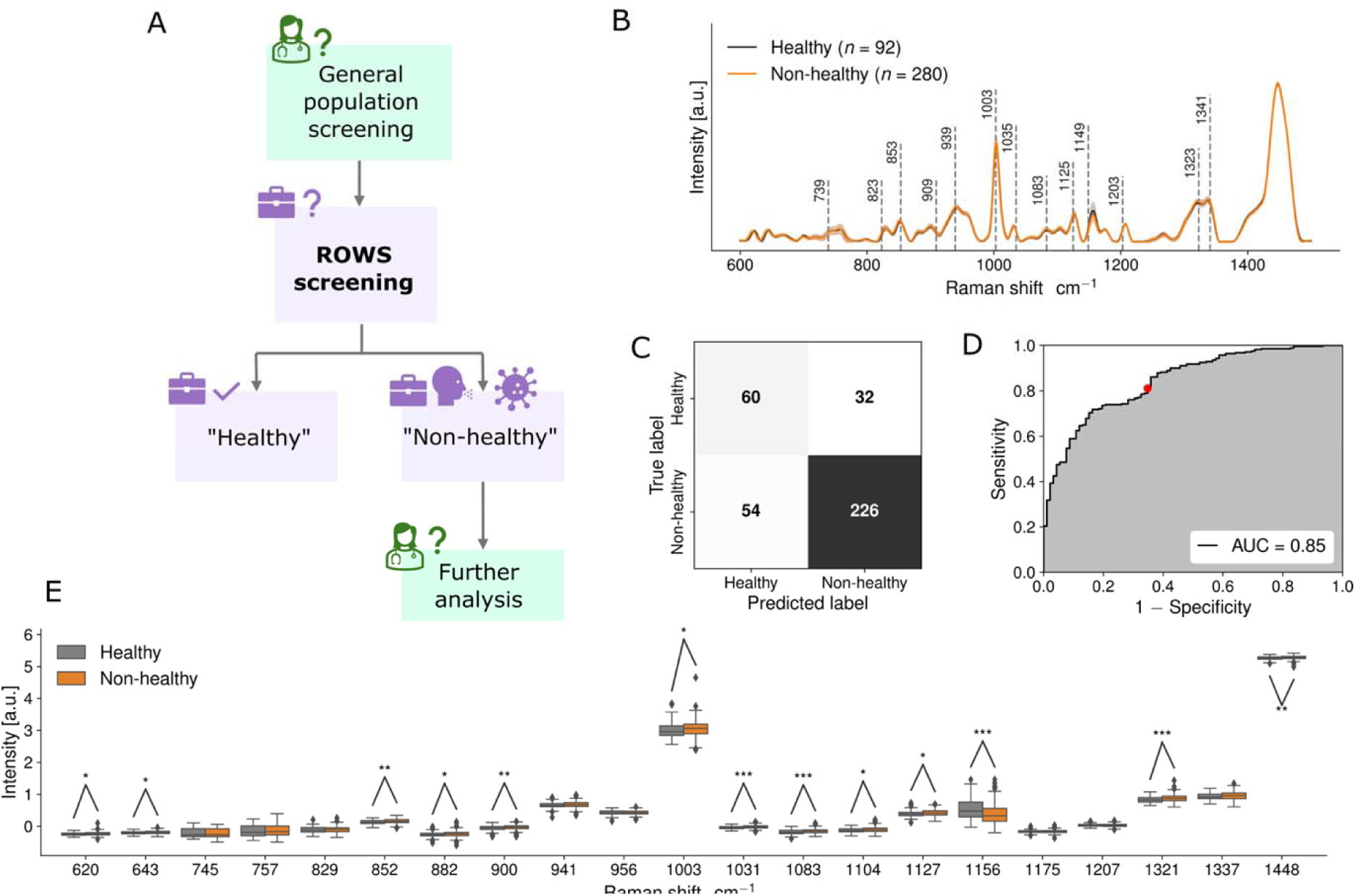
Towards point-of-care abnormal lung pathology screening assisted by Raman of well-based samples (Model 10): Raman spectroscopy detection of control vs. lung pathology, including benign tumors, asthma, and lung cancers. (A) Workflows of how a point-of-care ROWS device could be integrated into the clinical workflow. Green boxes indicate boxes where a highly trained clinician is required. Purple boxes indicate th screening and staging device outlined in this manuscript. (B) Raman spectra of liquid blood plasma from healthy controls (black line) and lung pathology (orange). Black dashed line show peaks used in the machine learning models. (B) Confusion matrix showing efficacy of Raman spectral prediction of controls and Stage I-IV lung cancer in liquid blood plasma. (C) Receiver operating characteristic (ROC) curves for machine learning models discriminating between liquid blood plasma spectral fingerprints from the two groups. (D) Box-and-whisker plot showing the relative intensities of different Raman bands between liquid blood plasma from control patients (grey) and those with Stage I-IV lung cancer (orange). * represents a p-value of ≤0.05. ** represents a p-value of ≤ 0.01. *** represents a p-value of ≤ 0.001.

### ROWS can discriminate between blood plasma samples from controls and non-cancer lung pathology patients

To investigate whether ROWS could potentially be used as a lung disease screening tool outside of the field of oncology, a machine learning model was created to discriminate between controls with no lung pathology and those with non-tumoral lung pathologies such as asthma, chronic obstructive pulmonary disease (COPD), interstitial lung disease or pulmonary arterial hypertension (PAH) (**Model 11, Figure 6**). This achieved an AUC of 0.96, an accuracy of 91%, a sensitivity of 91% and a specificity of 90%. Key Raman features included an increase in the peaks at 620, 852, 882, 900 and 941, 1031 and 1321 cm^-1^ which are primarily associated with protein, an increase in the peak due to phenylalanine, protein, and urea at 1003 cm^-1^, and an increase in peaks associated with lipids or fatty acids at 1083, 1104 and 1127 cm^-1^. There is a decrease in the features associated with certain protein/amino acid peaks at 745 and 757 cm^-1^ which was not seen in any of the machine learning models associated with lung cancer detection. Furthermore, there was no significant difference in the peak at 1156 cm^-1^ (carotenoids) which was present in all lung cancer detection models. This suggests that there are specific biomolecular changes in blood plasma associated with lung cancer which are different to those associated with other lung pathologies. There is an association between high blood urea nitrogen (BUN) to creatinine ratio in patients with COPD (*40*). This may account for the increase in the peak at 1003 cm^-1^. Serum analysis using non-Raman analysis shows that triglycerides, total cholesterol and LDL-cholesterol were elevated in patients with asthma which may account for the increase in peaks at 1083, 1104 and 1127 cm^-1^ (*41*). Circulating concentrations of carotenoids have been found to be lower in patients with both asthma and COPD (*42–45*). However, perhaps this reduction in carotenoid concentration is less pronounced than for lung cancer.

**Figure 6:**
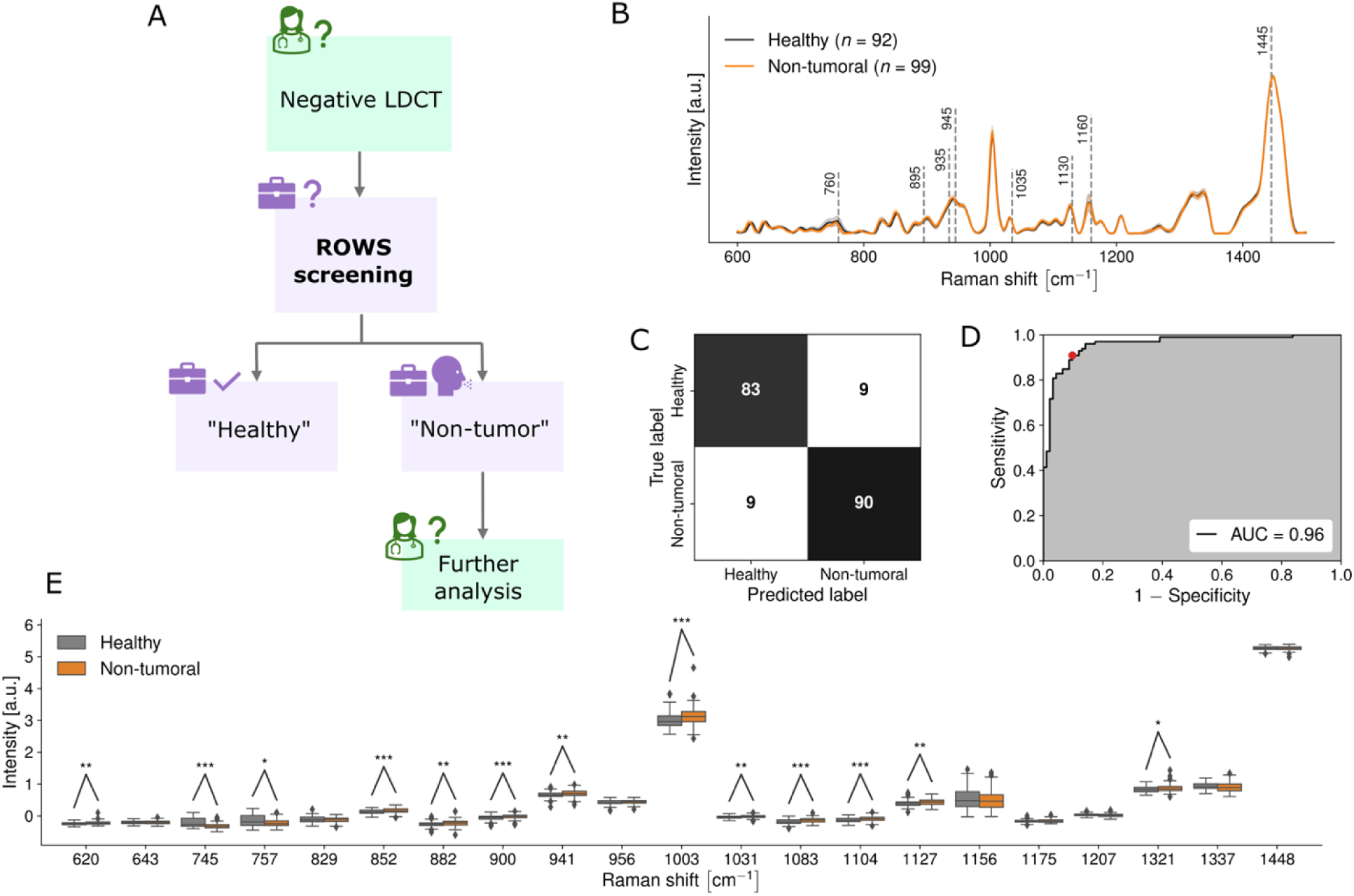
Towards point-of-care screening for non-cancer lung pathologies assisted by Raman of well-based samples (Model 11): Raman spectroscopy detection of controls vs. non-tumoral lung pathologies (e.g. asthma and chronic obstructive pulmonary disease). (A) Workflows of how a point-of-care ROWS device could be integrated into the clinical workflow. Green boxes indicate boxes where a highly trained clinician is required. Purple boxes indicate the screening and staging device outlined in this manuscript. (B) Raman spectra of liquid blood plasma from healthy controls (black line) and non-tumoral lung pathologies (orange). Black dashed lines show peaks used in the machine learning models. (B) Confusion matrix showing efficacy of Raman spectral prediction of healthy controls and non-tumoral lung pathologies in liquid blood plasma. (C) Receiver operating characteristic (ROC) curves for machine learning models discriminating between liquid blood plasma spectral fingerprints from the two groups. (D) Box-and-whisker plot showing the relative intensities of different Raman bands between liquid blood plasma from healthy control (grey) and those with non-tumoral lung pathologies (orange). * represents a p-value of ≤0.05. ** represents a p-value of ≤ 0.01. *** represents a p-value of ≤ 0.001.

ROWS could be used to assess low-risk symptomatic patients who had been for a one-off LDCT scan that was confirmed negative who were then sent for further assessment by a lung pathologist. Typically, the gold standard for screening of both asthma and COPD is spirometry. The sensitivity of spirometry is 23% and specificity is 90% for asthma,(*46*) and for COPD the sensitivity is 92% and specificity is 84% (*47*). ROWS could provide a more sensitive and specific alternative to spirometry.

### ROWS can discriminate between blood plasma samples from patients with benign lesions and lung cancer

Machine learning models built using ROWS measurements were able to discriminate between blood plasma samples from patients with benign lesions and lung cancer (**Models 12-14 and Supplementary Figures 9-11**). For these machine learning models, sensitivity and specificity were optimized in favor of high (>90%) specificity, to meet the requirements of LDCT-adjoint biofluid screening (*13*). Model 12, designed to discriminate between benign lesions and lung cancer regardless of stage, did so with 0.78 AUC, 65% accuracy, 59% sensitivity and 89% specificity (**Supplementary Figure 9, Table 2**). However, a small increase in specificity (92%) leads to a dramatic reduction in sensitivity (30%), suggesting that more samples from benign lesions are required to improve the machine learning accuracy. When sensitivity and specificity were balanced, accuracy was 72%, sensitivity was 72% and specificity was 78%. Peaks associated with proteins and lactic acid (852 and 941 cm^-1^), protein and lipid C-H bonds (882 and 900 cm^-1^) and protein amide III (1321 and 1337 cm^-1^) increased in cancer samples whilst the peak at 1448 cm^-1^ decreased (**Supplementary Figure 9D).** Model 13 classified benign lesions and early stage (I-II) cancers with a higher AUC of 0.82, accuracy of 67%, a sensitivity of 59% and a specificity of 92% (**Supplementary Figure 10, Table 2**). For the sensitivity-specificity balanced model, accuracy was 75%, sensitivity 73% and specificity 81%. Model 14 classified benign lesions and late stage (III-IV) cancers with an AUC of 0.94, accuracy of 88%, a sensitivity of 85% and a specificity 92% (**Supplementary Figure 11, Table 2**). For the sensitivity-specificity balanced model, accuracy was 88%, sensitivity 87% and specificity 89%. Model 14 likely outperforms Model 13 as later stage cancer is often associated with spread to lymph nodes,(*48*) metastasis, or growth affecting other organs or lung structures. This in turn can bring about more major changes in metabolism than for earlier stage cancers. Furthermore, later stage cancers were associated with an increase in circulating tumor cells (CTCs) which can impact the Raman spectrum of blood plasma. Model 12 is likely less accurate than Models 1 and 2 as the machine learning model must take into account the differences between the spectra between early-stage and late-stage cancers. The increase in accuracy with additional information about pathology (e.g. cancer stage, tumour type) has already been described in our Ember *et al.* 2024 paper concerning brain cancer.(*49*) Here, the generalized model for all brain tumors performed less well than the machine learning models trained for detection of specific types of brain tumor e.g. glioblastoma. There have not been published studies comparing serum metabolites from those with benign lesions and those with NSCLC. However, mass spectrometry was used to analyze metabolites showing differences between benign lesions and stage I lung adenocarcinoma, and found an increase in lactic acid which may account for the increase in peaks at 852 and 941 cm^-1^.(*50*)

The machine learning model for classification of blood plasma samples based on benign lesions or Stage I-IV lung cancer (**Model 13)** is currently insufficient for clinical incorporation due to the low number of benign samples. Further analysis of more benign samples would establish a more robust receiver operating characteristic (ROC) curve, and therefore allow sensitivity and specificity to be more accurately established.

To determine whether there were any biomolecular differences in plasma from non-tumoral pathology samples and patients with benign lesions, a machine learning model was created (**Model 15, Supplementary Figure 12**). The AUC was 0.95, accuracy 90%, sensitivity 89% and specificity 90%. There was a reduction in peaks associated with protein and amino acids (620, 882 cm^-1^), protein and lactate (852 cm^-1^), protein and lipid backbone (900, 941 cm^-1^), protein and urea (1003 cm^-1^) lipids and fatty acids (1104 cm^-1^), phosphate and citrate (956 cm^-1^) and carotenoids (1156 cm^-1^). There was an increase in peaks associated with protein and certain amino acids (745 and 757 cm^-1^) as well as protein and lipid C-H groups (1448 cm^-1^). Nine out of twelve of the key Raman peaks that were statistically different are the same as those in the machine learning model built to discriminate between healthy control and non-tumoral pathologies (Model 11), except the change is in the opposite direction.

To determine whether there were any biomolecular differences in plasma from healthy control samples and patients with benign lesions, machine learning Model 16 was created (**Model 16, Supplementary Figure 13**). The AUC was 0.83, accuracy 77%, sensitivity 75% and specificity 78%. Only five peaks were significantly different in intensity between the two groups: 757, 1031 and 1448 cm^-1^ which all were higher intensity in those with benign lesions and 956 and 1156 cm^-1^ which were lower in intensity.

Overall, the ROWS machine learning model for benign lesion detection with the highest accuracy was **Model 15**. This suggests that patients with benign lesions have biomolecular changes in their plasma that are of greater intensity when compared to patients with non-tumoral pathologies than when compared to healthy controls or patients with lung cancer.

### Smoking habits have little impact on the classification of samples by ROWS

To assess whether smoking habits affected the ability to detect lung cancer in blood plasma, machine learning models were built to include smoking as a confounding variable in ROWS measurements (**Models 17-20**, **Supplementary Figures 14-17**). This analysis showed that there was little impact on the classification of samples by ROWS.

## DISCUSSION

A lung pathology test that is low-cost, portable, user-friendly, minimally invasive, and accurate would have the potential to reduce lung cancer mortality. A Raman-based test could meet these criteria, improving the overall standard-of-care through more widespread screening in high-risk individuals and frequent testing for recurrence following treatment (**Figure 1**).

Previous groups have demonstrated the use of Raman spectroscopy in detecting lung cancer in serum or plasma outside of point-of-care settings. However, these studies have fallen short of the criteria for rapid, affordable, point-of-care analysis for several reasons: use of surface-enhanced Raman spectroscopy (SERS),(*51, 52*) dried samples,(*53*) microscope objectives(*51, 53–55*) and large sample volume.(*54*) The first limitation, SERS, uses metal nanoparticles or nanosurfaces to enhance the intrinsic Raman signal of nearby molecules. These increase test cost, and reduce reproducibility and user friendliness. (*56, 57*) The second limitation, sample drying, is time consuming: a 10 μl biofluid drop may take 45 minutes to dry. (*58*) Furthermore, blood spots dry heterogeneously, leading to intrasample variability in measurements if the laser spot size is smaller than the droplet.(*59*) The third limitation, microscope objectives may lead to susceptibility of spectra to the distance and position of the objective lens. This is because light is focused onto small volumes within the larger volume of the biofluid. This could feasibly lead to low signal-to-noise ratio through instrument or operator error. The fourth limitation is that some studies used blood product volumes greater than that provided by fingerprick-based blood extraction (100 μl or greater). This necessitates venous blood draws requiring clinically trained individuals, raising the cost and lowering the user-friendliness of the test. One final limitation is that in multiple studies, the stage of lung cancer was not detected.(*53*) This limits the clinical applicability of the tool.(*51, 54*)

Here, we show that different lung conditions have distinct profiles detected *via* a new technique: Raman of well-based samples (ROWS) combined with machine learning. This is achieved in 60 microlitres of liquid blood plasma in less than two minutes per sample. ROWS currently achieves accuracies of 72%-93% in detection of different lung conditions (**see Table 2**), paving the way for multiple clinical applications. ROWS allows classification of blood plasma samples based on whether they are from patients with Stage I-II or Stage III-IV lung cancers with 84% accuracy (**Model 1, Figure 2**). This staging could be invaluable for streamlining patient triaging, prognosis, and treatment plans. Furthermore, the ROWS system can detect a biomolecular signature associated with lung cancer amongst patients with lung pathologies with 93% accuracy and those without lung pathologies with 81% accuracy in a reagent-free manner (**Models 2 and 3, Figures 3 and 4**). Clinical use cases for **Models 2 and 3** could include screening of high-risk individuals or monitoring recurrence in previous lung cancer patients. It is important to note that lung cancer is relatively rare in the general population. Therefore, specificity would need to be extremely high for lung cancer screening in low-risk individuals. To achieve this use case, further studies with more biobanks must be carried out to provide larger, more diverse datasets for machine learning models. Nevertheless, the results presented here open the door to the eventual integration of ROWS into a rapid, point-of-care plasma-based screening tool for lung cancer detection in symptomatic patients or indeed a future population-wide lung cancer screening tool.

ROWS can also be used to detect abnormal lung pathology (including cancer and non-cancer pathologies) in an otherwise healthy population with 75% accuracy, which again could be used to help with triaging (**Model 10, Figure 5**). However, the accuracy is not high enough for a point-of-care diagnostic test for abnormal lung pathology and further investigations must be carried out to assess whether ROWS could be combined with symptoms to aid screening. Finally, ROWS can be used as a lung disease screening tool for non-cancer lung pathologies amongst the general population with 91% accuracy (**Model 11, Figure 6**). This could be used in patients with low lung cancer risk.

Raman spectroscopy and artificial intelligence can also be used to elucidate new cancer biomarkers. For example, there is a reduction in the peak due to carotenoids at 1156 cm^-1^ in detection of cancer compared to all negative groups (controls, non-tumoral pathologies, mixed negatives) except in the case of benign tumors. The concentration of carotenoids in blood plasma, or indeed the optical biomarker at 1156 cm^-1^ may therefore be used to select prospective patients for the Lung-RADS screening workflow.

All of the aforementioned machine learning models were achieved with a device costing less than USD 100,000, using a few tens of microlitres of plasma, in liquid form and without the addition of reagents or nanoparticles. Our Raman-based device addresses the challenge of low uptake of lung cancer screening with a minimally-invasive and low-cost solution. The device is also portable and user-friendly.

The next step towards point-of-care lung cancer detection device is integration of fingerpick analysis. This is not necessary for success of the device as venous blood draws are commonplace in a hospital setting. However, replacing venous blood draw with finger prick analysis would reduce the time needed from trained clinicians. The device will also be tested on a large scale in blood plasma samples fresh from patients who are undergoing lung cancer diagnosis, as in this study, all samples had been previously frozen.

It is important to note that the diagnostic performance reported in this study is likely a conservative estimate due to limited sample size. Each Raman spectrum contains over 1000 intensity values yet using all this data with the sample sizes in this study would result in model overfitting. To mitigate this risk, dimensionality reduction was implemented to constrict the feature space available to the machine learning models. This reduces model variance and improves generalization but also restricts the Raman data available to the machine learning models. It is therefore conceivable that the sensitivities and specificities reported here are a lower bounded estimate of the true potential for ROWS in lung cancer detection. To explore this, further analysis needs to be carried out with a larger sample size. As discussed, this is particularly important in the validation of whether ROWS could also be used to discriminate between benign lesions and malignant tumors. Another factor to consider is that these results were obtained with well-controlled study design in which confounding variables were minimized. Furthermore, there was a lack of diversity in age and ethnic diversity in all groups in the biobank (**Table 1**). For true clinical applicability, ROWS must be validated in a real-world setting and prove robust to sources of variance such as comorbidities, variations in sample collection and patient heterogeneity.

In conclusion, ROWS is a viable point-of-care lung pathology detection tool based on Raman spectroscopy and machine learning that can be used to detect multiple pathological factors using a small quantity of blood plasma. With more in-depth characterization, ROWS has the potential to revolutionize lung pathology testing, bringing it to a wider population, including remote and resource-limited settings.

## MATERIALS AND METHODS

### Sample Acquisition and Treatment

Three hundred and seventy-five blood plasma samples were analyzed from the UCPQ-ULaval Biobank, site of the AIRS Network Biobank (www.biobanque.ca/www.tissuebank.ca), in accordance with the management procedures approved by the research ethics committee. All participants provided written informed consent.

Clinical and pathological information from patient samples were collected (**Table 1)**. 51% of samples were from women and 49% from men. Samples were obtained from non-smokers, smokers, and ex-smokers. Lung cancer types included epidermal carcinoma, adenocarcinoma, and neuroendocrine tumors. Non-tumoral lung pathologies included asthma, chronic obstructive pulmonary disease (COPD), interstitial lung disease and pulmonary arterial hypertension (PAH). Blood plasma was obtained from volunteers as follows. For patients with lesions and lung cancer, samples were taken prior to lung resection for pulmonary lesions, and patients were not receiving any neoadjuvant therapy. This ensured that any observed differences were not due to surgery or neoadjuvant therapy. Controls were collected from individuals who had no lung pathologies but included those with asymptomatic hyperresponsiveness, allergic rhinitis without asthma, controls without asthma and without allergies, allergies without asthma or rhinitis. Plasma was collected in 4ml K2-EDTA BD Vacutainers with mauve stoppers. Blood was centrifuged immediately or conserved at 4°C for a maximum of 3 hours before centrifugation. Centrifugation was carried out at 3200g at room temperature for 15 min, and the upper layer (plasma) was taken and pipetted into 200 µl aliquots, before freezing at -80 °C.

### Sample Analysis

Samples were thawed at room temperature for approximately 22 minutes and vortexed for 30 seconds. Samples were analyzed with a custom, portable, rapid single-point Raman spectrometer in the form of a suitcase. The system uses a 785 nm laser source (Model IO785MM1500M4S, Innovative Photonic Solutions, USA) with an output of 1.5 W and a spectral bandwidth < 2 nm.(*60, 61*) The other main constituent of the system is a spectrometer (HT model, EmVision, USA) composed of a diffraction grating and a charge-coupled device (CCD) camera (Newton 920, Oxford Instruments, USA) resulting in a spectral resolution < 8.7 cm^-1^.

Before each measurement the system CCD sensor was cooled to -80°C. Calibration of the *x*-axis (Raman shift) was determined from a spectrum acquired using acetaminophen powder prior to each measurement. The system response across the spectral domain of detection was determined using the fluorescence spectrum of a standard reference material (SRM 2214, National Institute of Standards and Technology, NIST, USA). These measurements were used to normalize each plasma measurement allowing interpretation of the spectra at a biochemical level through identification of peaks.

For each blood plasma spectrum, a 50 µl sample was analyzed after being pipetted into an aluminum well in a sample cassette. The cassette was constructed from a 3D printed plastic rectangular strip of plastic with a small hole in one end. Aluminum tape was used to create the well itself. Spectra were obtained using a laser power of 861 mW at the sample, with 3 measurements per drop. Automatic exposure control -where sensor exposure time is automatically adjusted to maximize dynamical range usage- was implemented, with an average exposure time of 1.2 +/- 0.3 s, a minimum exposure time of 380 ms and a maximum exposure time of 3000 ms. Fifty accumulations were obtained per sample, but only the first 20 accumulations were required and used in the analysis. The laser spot size had an on-sample diameter of x mm, resulting -because of light diffusion effects and reflections on the lower level of the cuvette- in a measurement effectively interrogating the whole volume of the sample.

### Data processing

A number of data processing steps were required to extract a Raman spectrum out of a raw unprocessed acquisition.(*62*) In the following order, the steps were 1) background (dark noise) subtraction obtained with the laser turned off, 2) removal of cosmic ray events, 3) *x*-axis calibration by corresponding Raman peaks from Tylenol measurements to their known position in cm^-1^, in steps of 1 cm^-1^, 4) correction by the instrument response function (*y*-axis calibration) obtained from a calibration material (NIST 785 nm Raman standard), 5) truncation of the pixels below 450 cm^-1^, 6) averaging of successive accumulations performed on the same sample, 7) baseline removal using the BubbleFill algorithm with adjusted minimal bubble width, 8) further constrain the final Raman spectrum between 600 and 1500 cm^-1^ by truncation, to remove the water peak present at higher wavenumbers which can vary based on patient hydration or sample drying, and 9) standard normal variate (SNV) normalization. All spectra are individually interpolated between 600 and 1500 cm^-1^ in steps of 1 cm^-1^.

### Classification models

In total 20 machine learning models were produced to assess the degree of separability between different subgroups, in terms of biomolecular features extracted from plasma Raman spectra (**Table 2**). A two-step approach for this to ensure molecular interpretability of the predictive models. First, a dimensional reduction step (feature selection) led to the identification of a subset of individual spectral features -in the form of inelastic scattering intensities at specific wavenumber values- that had the most discriminative power between the classes, for each of the 20 models. Secondly, those dimensionally reduced sets of features were used to to train and validation the machine learning models. The pre-selection of a finite set of features was required to minimize the likelihood that models led to overfitting of the data and thus allowed predictive performances to be reported that were realistic and reflective of real-world scenarios.

To ensure classification results were derived from Raman bands rather than noise, we applied a feature reduction procedure. Gaussians were fitted to any peak with an SNV-normalized intensity of higher than 0.35 to extract peak height, width and position (with 0.2 cm^-1^ tolerance to position). Only the peaks present in at least half of samples were considered, to reduce risk of artifacts. The result is a set of 21 Raman bands (Fig. 2) for which we constrained the spectral bins to the full width at half maximum of each band, such that the total number of features (spectral bins) is thus reduced from 900 to 359.

Each classification model was trained following a 5-fold cross validation to assess the model performance. To do so, the dataset was split into 5 subsets. Four of these subsets (training set) were used to train a model which was then applied on the remaining set (validation set) to evaluate the performance. This was repeated until each subset had been used once as a validation set. Independently for each fold, the feature selection algorithm (a support vector machine (SVM) with a linear kernel and L1 regularization) was be applied on the reduced feature set described above, followed by the classification algorithm (a SVM with linear kernel with L2 regularization).

Specific parameters of the feature selection and classification algorithms needed to be adjusted, which was done via a grid search running over multiple combinations. The regularization parameter *C* for the feature selection, was varied between 0.05 and 0.5 and the maximum number of features between 10 and 30. The regularization parameter *C* for the classification algorithm ranged between 0.01 and 1. For each combination, the performance was assessed via the 5-fold cross validation described above, where predictions on the validation sets are compared with the known label associated with each sample. The prediction probabilities on the validation set were used to compute a receiver operating characteristic (ROC) curve. The ROC curve gave the area under curve (AUC) –with an optimal value of 1 for perfect separation between classes– but also enabled to extract a threshold that optimized specificity and sensitivity of detection These values were used to turn prediction probabilities into binary predictions to make a confusion matrix (CM). The confusion matrices provided the number of false/true positives and false/true negatives. The combination of ROC curve AUC and CM values was used to select the bet set of parameters (*C*, number of features) and thus the best classification model, which we report results on. For each model, p-values for the 21 Raman bands were computed using the Wilcoxon-Mann-Whitney test and are reported as box and whisker plots. Twenty machine learning models (**Table 2)** were tested. Unless specified, sensitivity and specificity were calculated from the point at the top left corner of the ROC curve, optimizing for both sensitivity and specificity.

### Biomolecular peak assignment

As a single species of biomolecule can give rise to multiple Raman peaks, correlation analysis was undertaken to examine the interrelatedness of different peaks within the blood plasma Raman spectra. To measure interrelatedness of Raman peaks, the change in mean peak intensity for nineteen peaks of interest was evaluated for each machine learning model (**Supplementary Figure 1**). Two out of the original twenty-one peaks (1175 and 1207 cm^-1^) showed no statistically different change in any machine learning model, so these were left out of the analysis. For each peak, the change in mean peak intensity was represented in **Supplementary Figure 1** as one of the following: not statistically different (blank), statistically greater in the positive than the negative group (up) or statistically less in the positive than the negative group (down). To generate a numerical value for interrelatedness of a pair of peaks, correlation values were calculated for each machine learning model. If at least one of the two peaks in the pair had no statistical change in mean peak intensity, the peaks were considered non-correlated (a value of 0). If the change in mean peak intensity matched for a given machine learning model (e.g. if both peaks were higher in the positive group than the negative group), the peaks were considered correlated for that machine learning model, and a value of +1 was given. If the change in mean peak intensity was in opposing directions for a particular machine learning model, a value of -1 was given.

The correlation values for all machine learning models for a pair of peaks were then summed to give the total correlation value for that pair. This is represented as a confusion matrix (**Supplementary Figure 2**). Using this confusion matrix and spectra from literature, biomolecular assignments were made (**Supplementary Table 3**).

## List of Supplementary Materials

Supplementary Tables 1-3

Supplementary Figures 1-17

Smoking Status

## Acknowledgements

The authors would like to thank the team at the IUCPQ-ULaval site of the AIRS Network Tissue Biobank for their valuable assistance, in particular Dr Philippe Joubert and Sabrina Biardel.

## Funding

This research was funded by a National Science and Engineering Research Council (NSERC) Alliance Grant in collaboration with Reveal Life Science (previously Exclaro-Tridan) (FL).

## Author contributions

F.D.1 = Frederick Dallaire, F.D.2 = Francois Daoust Conceptualization: K.E. and F.L. Methodology: K.E., F.D.1., F. D.2, N.K., G.S., R.L.R, F. L. Ethical approval and biobank access: J.S. Device software: G.S. Device design: N.K. Measurement acquisition: E.D., M.B. Biochemical assignments: K.E. Resources: F.L. Data curation: K.E., F.D.1 Machine learning: F.D.1, G.S. Data analysis: K.E., F.D. Figures: K.E., F.D. Writing—original draft: K.E., F.D.1 Writing—review and editing: K.E., F. D.1, M.L., F.L. Supervision: F.L. Funding acquisition: K.E. and F.L.

## Competing Interests

Frédéric Leblond, Francois Daoust and Nassim Ksantini are shareholders of Reveal Life Science. Francois Daoust, Juliette Selb and Nassim Ksantini are employees of Reveal Life Science.

## Data and materials availability

Data cannot be shared due to patient privacy and agreements with the AIRS Network Tissue Biobank. Data processing code is available through Github. Machine learning models are described in detail in Ember *et al.* Scientific Reports 2024. Code repository for model training, analysis and validation is publicly available in the paper “Open-sourced Raman spectroscopy data processing package implementing a novel baseline removal algorithm validated from multiple datasets acquired in human tissue and biofluids” Sheehy et al., Journal of Biomedical Optics, (2023) and also on Github (https://github.com/mr-sheg/orpl).

## Notes

### Competing Interest Statement

Frederic Leblond, Francois Daoust and Nassim Ksantini are shareholders of Reveal Life Science. Francois Daoust, Juliette Selb and Nassim Ksantini are employees of Reveal Life Science.

### Author Declarations

The Research Ethics Board of the Centre Hospitalier de l'universite de Montreal (CHUM) gave ethical approval for this work.

