## Supplementary Materials for "A method for lung cancer detection and staging from a drop of blood plasma via Raman spectroscopy of well-based samples (ROWS)"

**Supplementary figures and tables**

**Supplementary Table 1: Machine learning models developed using Raman spectra of blood plasma from a biobank acquired from lung cancer patients, healthy controls and patients with other lung pathologies for individuals with different smoking status.**

| **Model 17** | 69 | Non-tumoral non-smoker | 27 | Non-tumoral smoker |
| --- | --- | --- | --- | --- |
| **Model 18** | 41 | Healthy non-smoker | 48 | Healthy ex-smoker |
| **Model 19** | 43 | Stage I-II ex-smoker | 48 | Stage I-II smoker |
| **Model 20** | 48 | Healthy ex-smoker | 38 | Stage III-IV ex-smoker |

**Supplementary Table 2: Performance metrics of the machine learning models based on smoking status developed using Raman spectra of blood plasma from a biobank acquired from lung cancer patients, healthy controls and patients with other lung pathologies**

| **Model 17** | 74 | 74 | 74 | 0.78 |
| --- | --- | --- | --- | --- |
| **Model 18** | 69 | 75 | 61 | 0.66 |
| **Model 19** | 58 | 50 | 67 | 0.56 |
| **Model 20** | 81 | 79 | 83 | 0.90 |

| **Peak (cm^-1^)** | **M1** | **M2** | **M3** | **M4** | **M5** | **M6** | **M7** | **M8** | **M9** | **M10** | **M11** | **M12** | **M13** | **M14** | **M15** | **M16** | **M17** | **M18** | **M19** | **M20** |
| --- | --- | --- | --- | --- | --- | --- | --- | --- | --- | --- | --- | --- | --- | --- | --- | --- | --- | --- | --- | --- |
| 620 |  |  | up |  |  | up |  |  |  | up | up |  |  |  | down |  |  |  | down |  |
| 643 | up | up | up |  | up | up | up |  | up | up |  |  |  | up |  |  | up |  |  | up |
| 745 | up | up | up |  | up | up | up | up | up |  | down |  |  |  | up |  |  |  |  | up |
| 757 | up | up | up |  | up | up | up |  | up |  | down |  |  | up | up | up |  |  |  | up |
| 829 | down | up | up | up |  | up | up | up |  |  |  |  | up |  |  |  |  |  |  |  |
| 852 | down |  | up | up |  | up | up | up |  | up | up | up | up |  | down |  |  |  |  |  |
| 882 | up |  | up |  | up | up |  |  | up | up | up | up |  | up | down |  |  |  | down | up |
| 900 |  |  | up |  | up | up |  |  |  | up | up | up | up | up | down |  |  |  |  | up |
| 941 | down | down |  | up |  | up |  |  |  |  | up | up | up |  | down |  |  |  |  |  |
| 956 |  | down |  |  |  |  |  |  |  |  |  |  | up |  | down | down |  |  |  |  |
| 1003 | down | down |  | up |  |  |  |  | down | up | up |  |  |  | down |  |  |  |  |  |
| 1031 |  |  | up | up | up |  |  |  |  | up | up |  |  |  |  | up |  |  |  | up |
| 1083 | down | down | up | up |  | up |  |  | down | up | up |  |  |  |  |  |  |  |  |  |
| 1104 | down | down |  | up | down |  |  | up | down | up | up |  | up | down | down |  |  |  |  | down |
| 1127 | down |  |  | up |  |  |  |  | down | up | up |  |  |  |  |  |  |  |  |  |
| 1156 |  | down | down | down | down | down | down | down | down | down |  |  |  |  | down | down | down |  |  | down |
| 1321 | down | up | up | up | up | up | up | up |  | up | up | up | up |  |  |  |  | down |  | up |
| 1337 |  | up | up | up | up | up | up | up | up |  |  | up |  | up |  |  |  | down |  | up |
| 1448 | up | up | up |  | up |  | up |  | up | up |  | down | down |  | up | up |  |  |  | up |

**Supplementary Figure 1: Response of different Raman peaks in machine learning models.** The change in mean peak intensity is represented in **Supplementary Figure 3** as one of the following: not statistically different (blank), statistically greater in the positive than the negative group (up) or statistically less in the positive than the negative group (down).

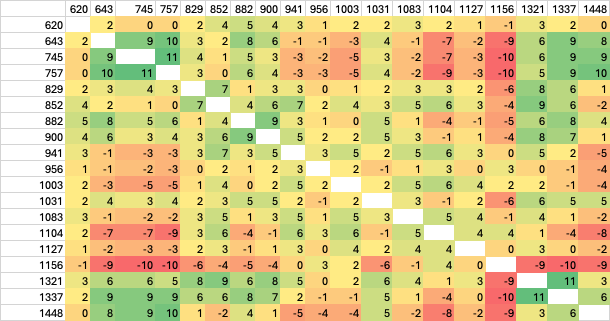

**Supplementary Figure 2: Confusion matrix of Raman features used in machine learning models.** Higher numbers and green boxes indicate higher degrees of correlation between peaks. Lower numbers and red boxes indicate lower degrees of correlation between peaks. Degree of correlation was used to inform biomolecular assignments in Supplementary Table 3.

**Supplementary Table 3: Biomolecular assignments for Raman features in Raman spectra of blood plasma.**

| **Raman peak (wavenumbers)** | **Biomolecular assignment** |
| --- | --- |
| 620 | Amino acids (phenylalanine, glutamate)^38^ |
| 643 | Protein (tyrosine)^36,38^ |
| 745 | Protein (tryptophan, phenylalanine, aspartate, isoleucine) ^36,38^ |
| 757 | Protein (tryptophan, phenylalanine, aspartate, isoleucine) ^36,38^ |
| 829 | Lactic acid,^39^ citrate,^40^ protein^36^ |
| 852 | Glycerol, ^41^ protein (tyrosine, alanine, leucine, lysine, proline) ^36,38^ |
| 882 | Protein (tryptophan),^36^ phosphate |
| 900 | Protein (C-H),^36^ lipid (C-H)^37^ |
| 941 | Protein (N-C-C),^36^ lipids backbone^37^ |
| 956 | Phosphate, citrate^40^ |
| 1003 | Protein (phenylalanine, tryptophan),^36^ urea ^42^01/12/2025 17:08:00 |
| 1031 | Protein (phenylalanine, arginine, glycine, lysine, isoleucine) ^36,38^ |
| 1083 | Fatty acid (C-C), lipids^37^ |
| 1104 | Fatty acid (C-C), lipids^37^ |
| 1127 | Fatty acid (C-C), lipids,^43^ protein (C-N, serine),^36,38^ glucose2025-12-01 5:08:00 PM |
| 1156 | Carotenoids^44^, fatty acids (C-C)^37^ |
| 1321 | Protein (C-H, amide III)^36^ |
| 1337 | Protein (C-H, aspartate, histidine, proline, valine, tryptophan) ^36,38^ |
| 1448 | Protein (C-H, isoleucine, valine),^36,38^ lipid (CH_2_/CH_3_)^37^ |

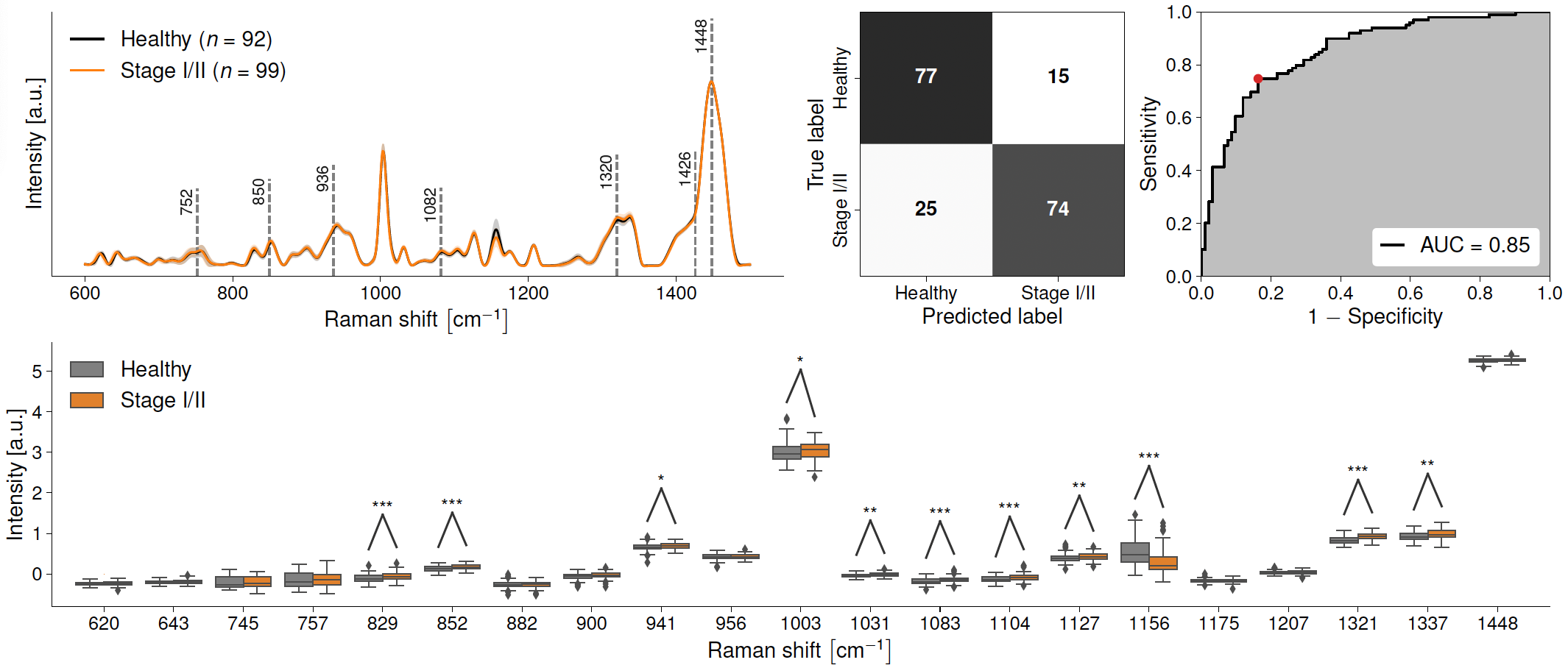

**Supplementary Figure 3: Raman spectroscopy detection of healthy control vs. Stage I-II lung cancers (Model 4).** (A) Raman spectra of liquid blood plasma from healthy controls (black line) and Stage I-II lung cancer (orange). Black dashed lines show peaks used in the machine learning models. (B) Confusion matrix showing efficacy of Raman spectral prediction of controls and Stage I-II lung cancer in liquid blood plasma. (C) Receiver operating characteristic (ROC) curves for machine learning models discriminating between liquid blood plasma spectral fingerprints from the two groups. (D) Box-and-whisker plot showing the relative intensities of different Raman bands between liquid blood plasma from control patients (grey) and those with Stage I-II lung cancer (orange). * represents a p-value of ≤0.05. ** represents a p-value of ≤ 0.01. *** represents a p-value of ≤ 0.001.

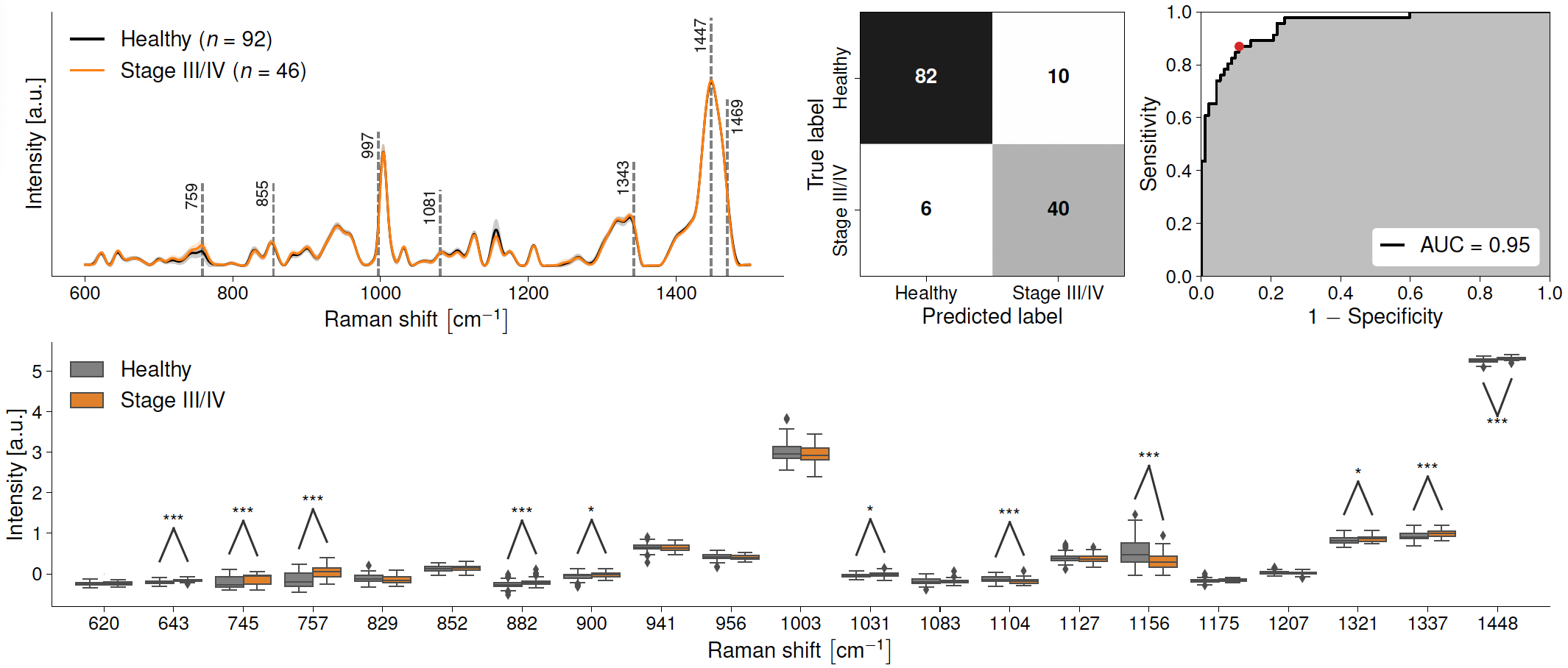

**Supplementary Figure 4: Raman spectroscopy detection of healthy control vs. Stage III-IV lung cancers (Model 5).** (A) Raman spectra of liquid blood plasma from healthy controls (black line) and Stage III-IV lung cancer (orange). Black dashed lines show peaks used in the machine learning models. (B) Confusion matrix showing efficacy of Raman spectral prediction of controls and Stage III-IV lung cancer in liquid blood plasma. (C) Receiver operating characteristic (ROC) curves for machine learning models discriminating between liquid blood plasma spectral fingerprints from the two groups. (D) Box-and-whisker plot showing the relative intensities of different Raman bands between liquid blood plasma from control patients (grey) and those with Stage III-IV lung cancer (orange). * represents a p-value of ≤0.05. ** represents a p-value of ≤ 0.01. *** represents a p-value of ≤ 0.001.

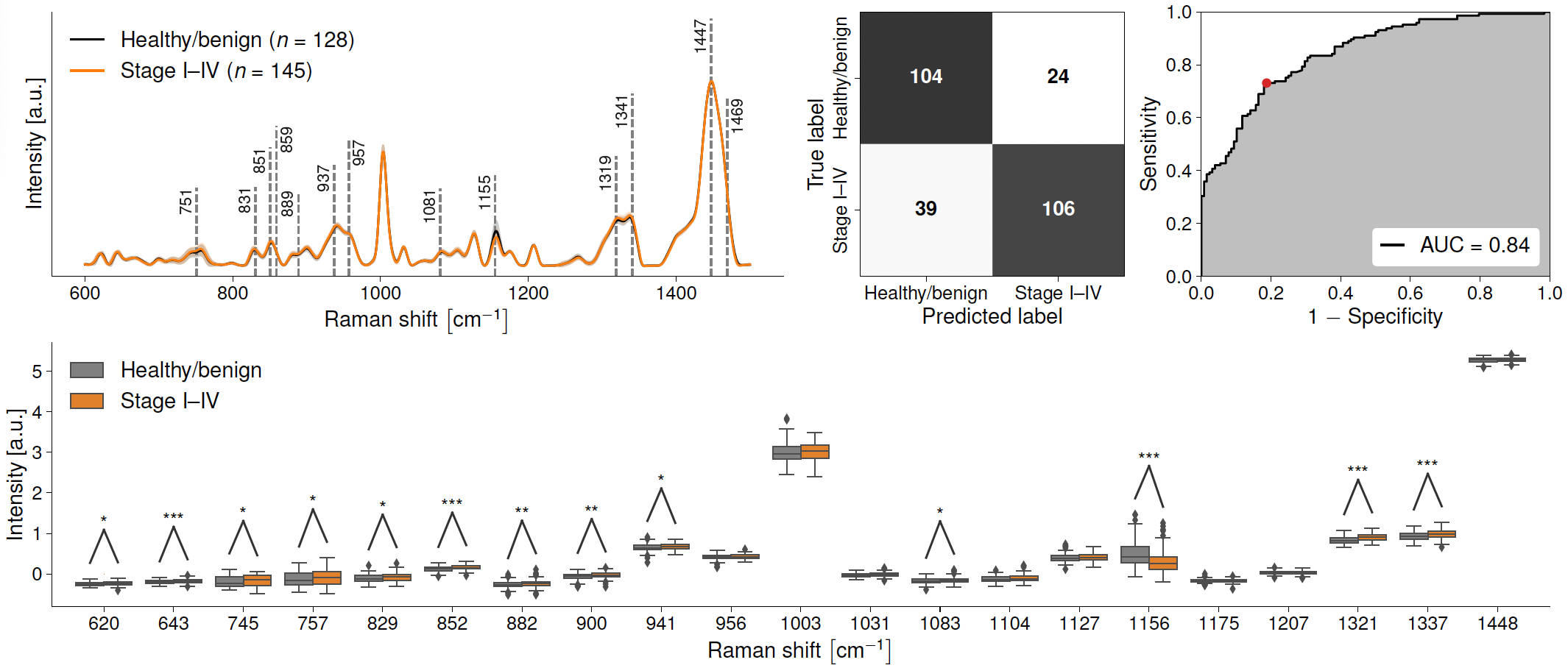

**Supplementary Figure 5: Raman spectroscopy detection of healthy controls and benign tumors vs. Stage I-IV lung cancers (Model 6).** (A) Raman spectra of liquid blood plasma from healthy controls and benign tumors (black line) and Stage I-IV lung cancer (orange). Black dashed lines show peaks used in the machine learning models. (B) Confusion matrix showing efficacy of Raman spectral prediction of controls/benign tumors and Stage I-IV lung cancer in liquid blood plasma. (C) Receiver operating characteristic (ROC) curves for machine learning models discriminating between liquid blood plasma spectral fingerprints from the two groups. (D) Box-and-whisker plot showing the relative intensities of different Raman bands between liquid blood plasma from control/ benign tumor patients (grey) and those with Stage I-IV lung cancer (orange). * represents a p-value of ≤0.05. ** represents a p-value of ≤ 0.01. *** represents a p-value of ≤ 0.001.

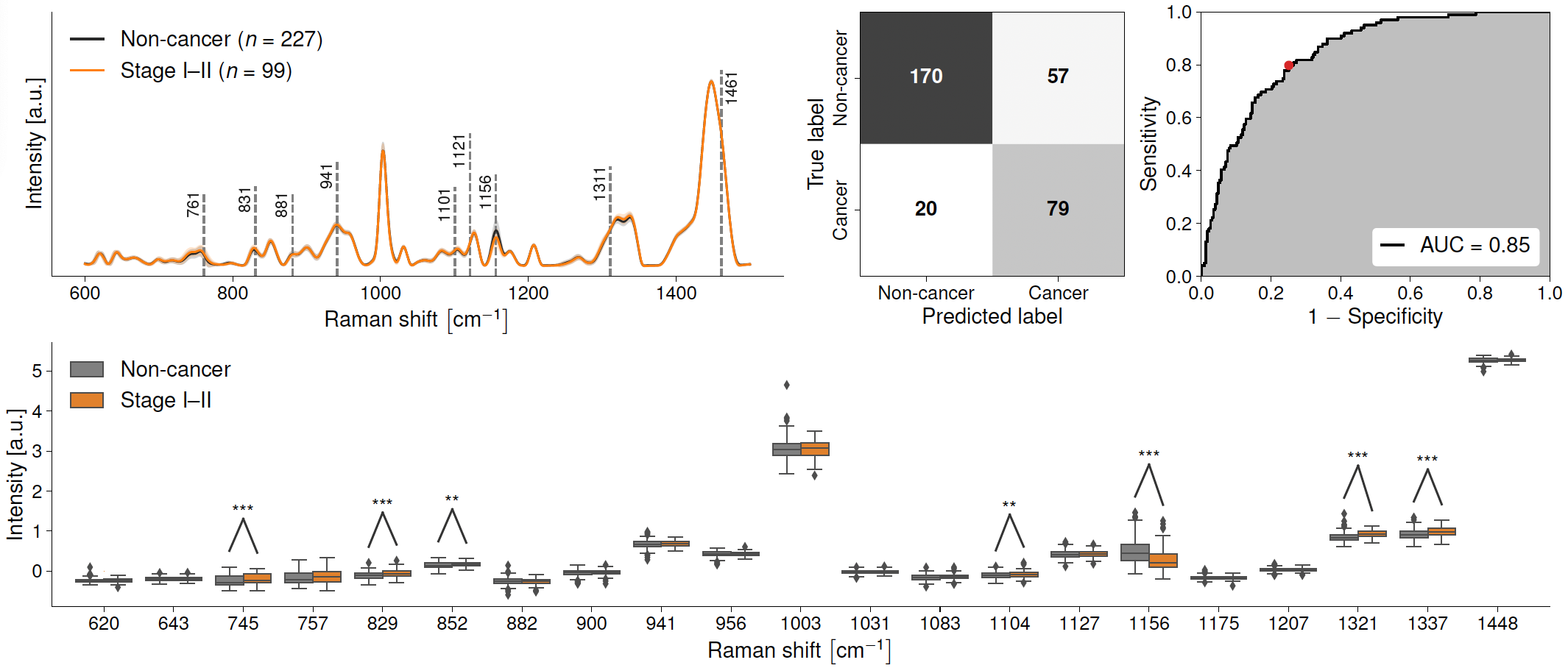

**Supplementary Figure 6: Raman spectroscopy detection of non-cancer samples vs. Stage I-II lung cancer (Model 7).** (A) Raman spectra of liquid blood plasma from non-cancer (black line) and Stage I-II cancer (orange). Black dashed lines show peaks used in the machine learning models. (B) Confusion matrix showing efficacy of Raman spectral prediction of non-cancer and Stage I-II cancer in liquid blood plasma. (C) Receiver operating characteristic (ROC) curves for machine learning models discriminating between liquid blood plasma spectral fingerprints from the two groups. (D) Box-and-whisker plot showing the relative intensities of different Raman bands between liquid blood plasma from non-cancer (grey) and those with Stage I-II cancer (orange). * represents a p-value of ≤0.05. ** represents a p-value of ≤ 0.01. *** represents a p-value of ≤ 0.001.

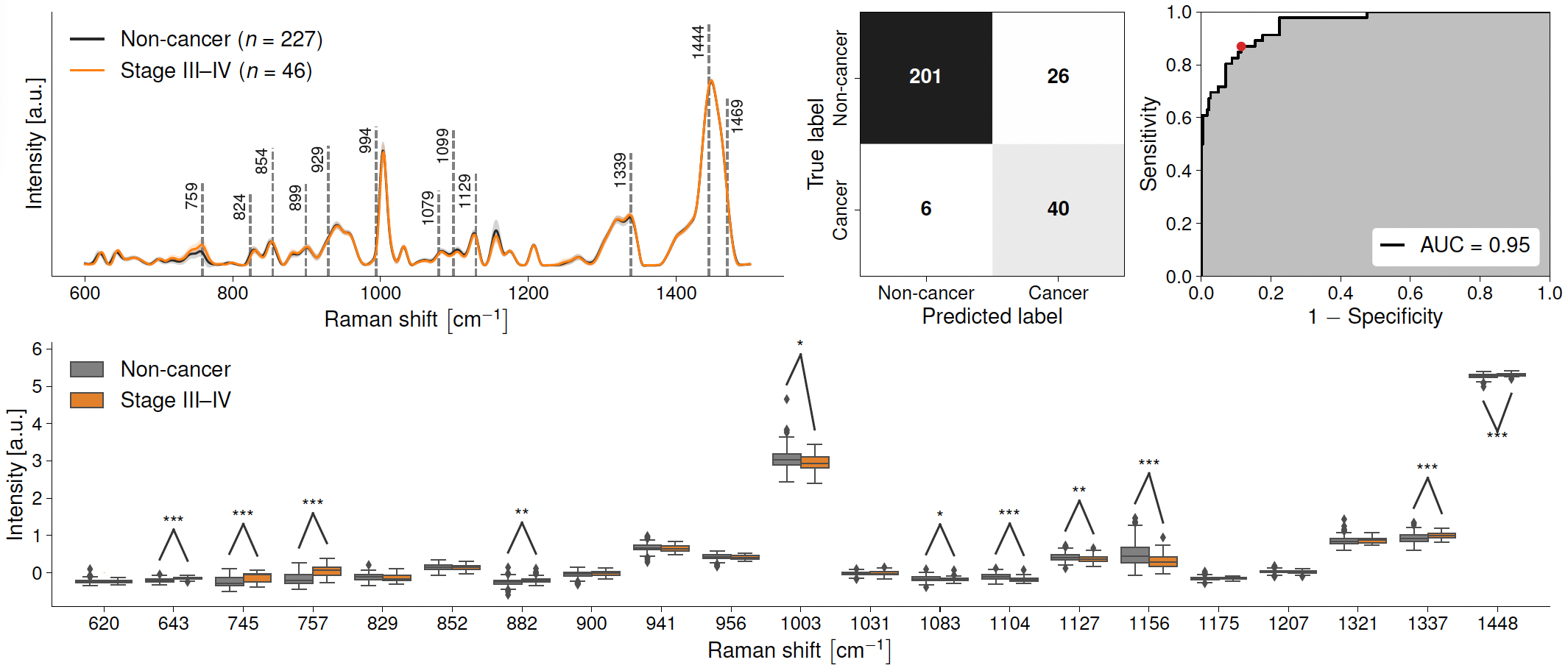

**Supplementary Figure 7: Raman spectroscopy detection of non-cancer samples vs. Stage III-IV lung cancer (Model 8).** (A) Raman spectra of liquid blood plasma from non-cancer (black line) and Stage III-IV cancer (orange). Black dashed lines show peaks used in the machine learning models. (B) Confusion matrix showing efficacy of Raman spectral prediction of non-cancer and Stage III-IV cancer in liquid blood plasma. (C) Receiver operating characteristic (ROC) curves for machine learning models discriminating between liquid blood plasma spectral fingerprints from the two groups. (D) Box-and-whisker plot showing the relative intensities of different Raman bands between liquid blood plasma from non-cancer (grey) and those with Stage III-IV cancer (orange). * represents a p-value of ≤0.05. ** represents a p-value of ≤ 0.01. *** represents a p-value of ≤ 0.001.

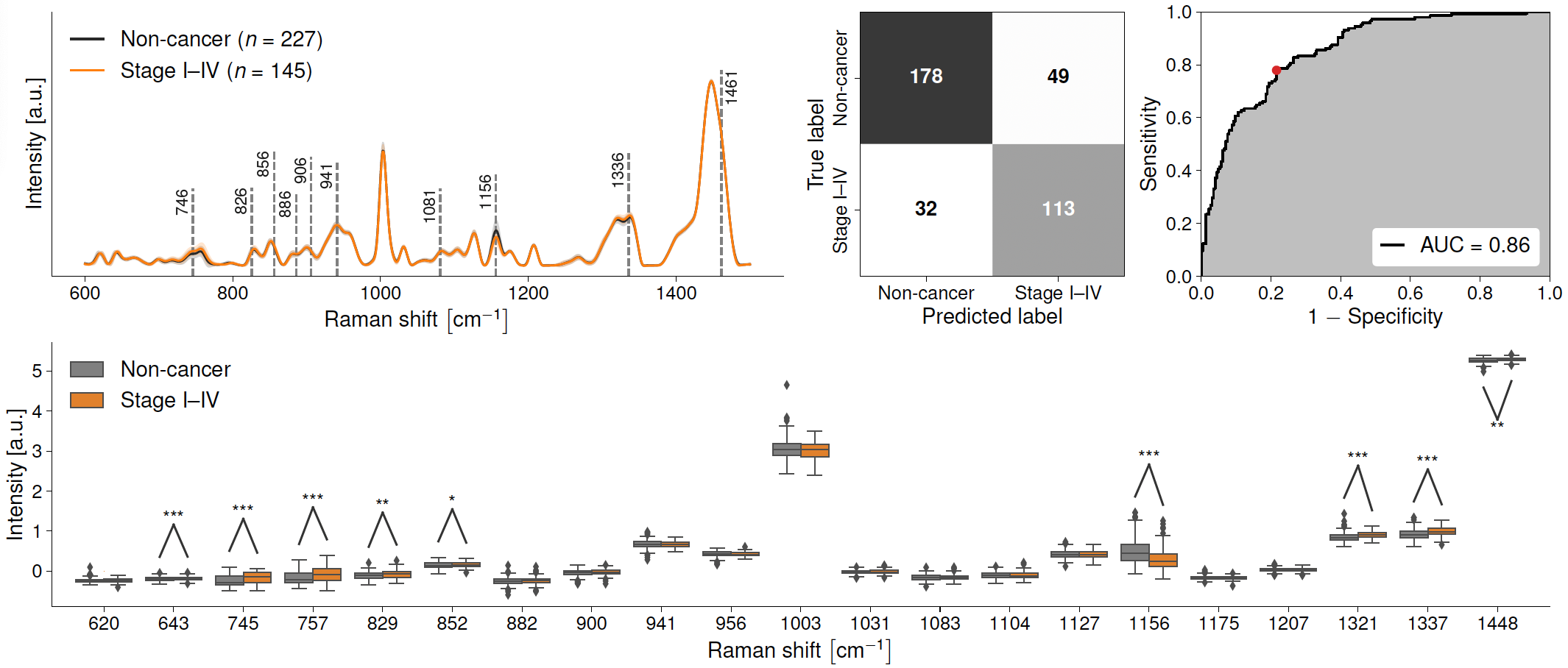

**Supplementary Figure 8: Raman spectroscopy detection of non-cancer samples vs. Stage I-IV lung cancer (Model 9).** (A) Raman spectra of liquid blood plasma from non-cancer (black line) and Stage I-IV cancer (orange). Black dashed lines show peaks used in the machine learning models. (B) Confusion matrix showing efficacy of Raman spectral prediction of non-cancer and Stage I-IV cancer in liquid blood plasma. (C) Receiver operating characteristic (ROC) curves for machine learning models discriminating between liquid blood plasma spectral fingerprints from the two groups. (D) Box-and-whisker plot showing the relative intensities of different Raman bands between liquid blood plasma from non-cancer (grey) and those with Stage I-IV cancer (orange). * represents a p-value of ≤0.05. ** represents a p-value of ≤ 0.01. *** represents a p-value of ≤ 0.001.

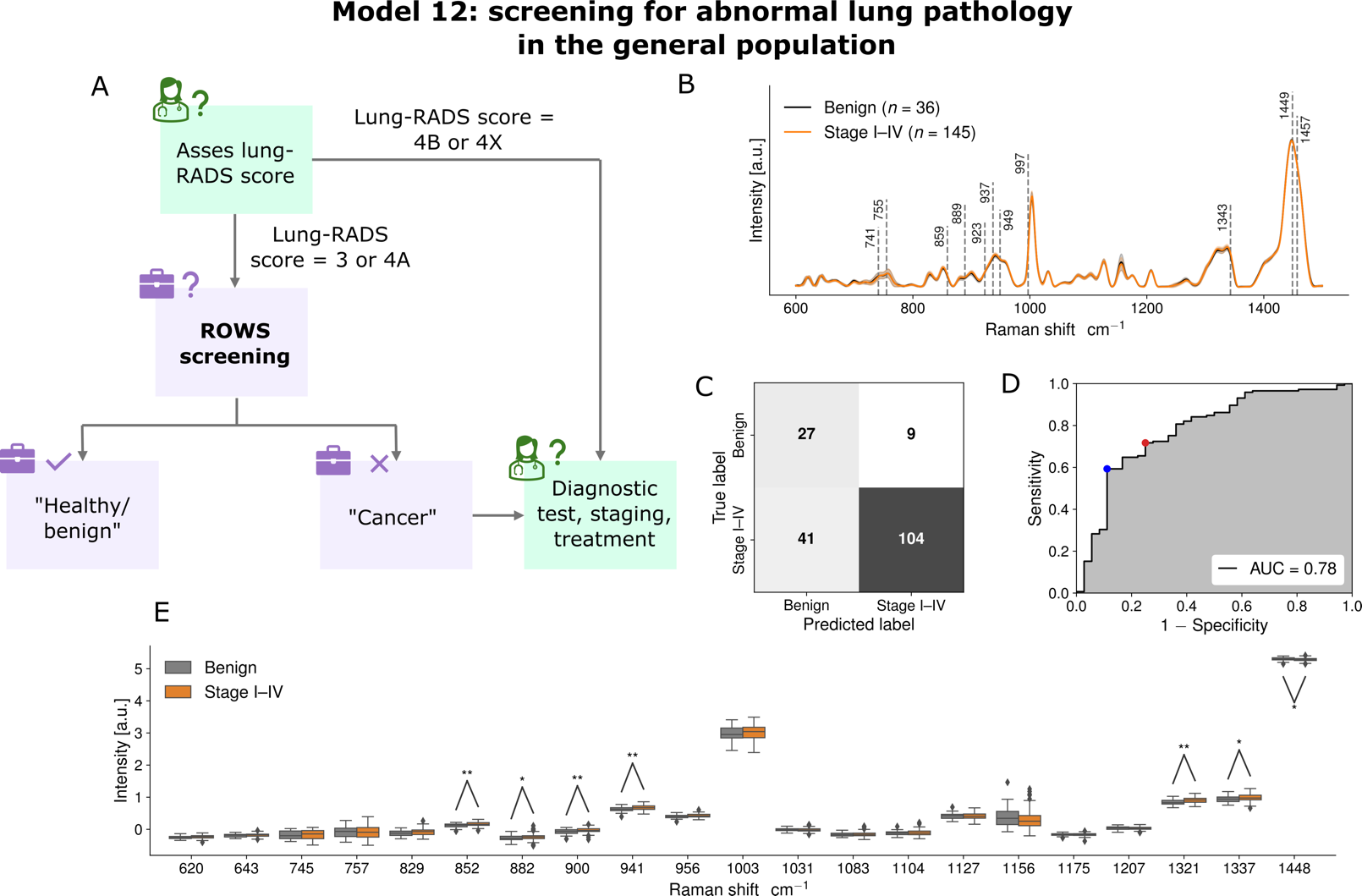

**Supplementary Figure 9: Raman spectroscopy detection of benign tumors vs. Stage I-IV lung cancers (Model 12).** (A) Raman spectra of liquid blood plasma from patients with benign tumors (black line) and Stage I-IV lung cancer (orange). Black dashed lines show peaks used in the machine learning models. (B) Confusion matrix showing efficacy of Raman spectral prediction of benign tumors and Stage I-IV lung cancer in liquid blood plasma. (C) Receiver operating characteristic (ROC) curves for machine learning models discriminating between liquid blood plasma spectral fingerprints from the two groups. (D) Box-and-whisker plot showing the relative intensities of different Raman bands between liquid blood plasma from patients with benign tumors (grey) and Stage III-IV lung cancer (orange). * represents a p-value of ≤0.05. ** represents a p-value of ≤ 0.01. *** represents a p-value of ≤ 0.001.

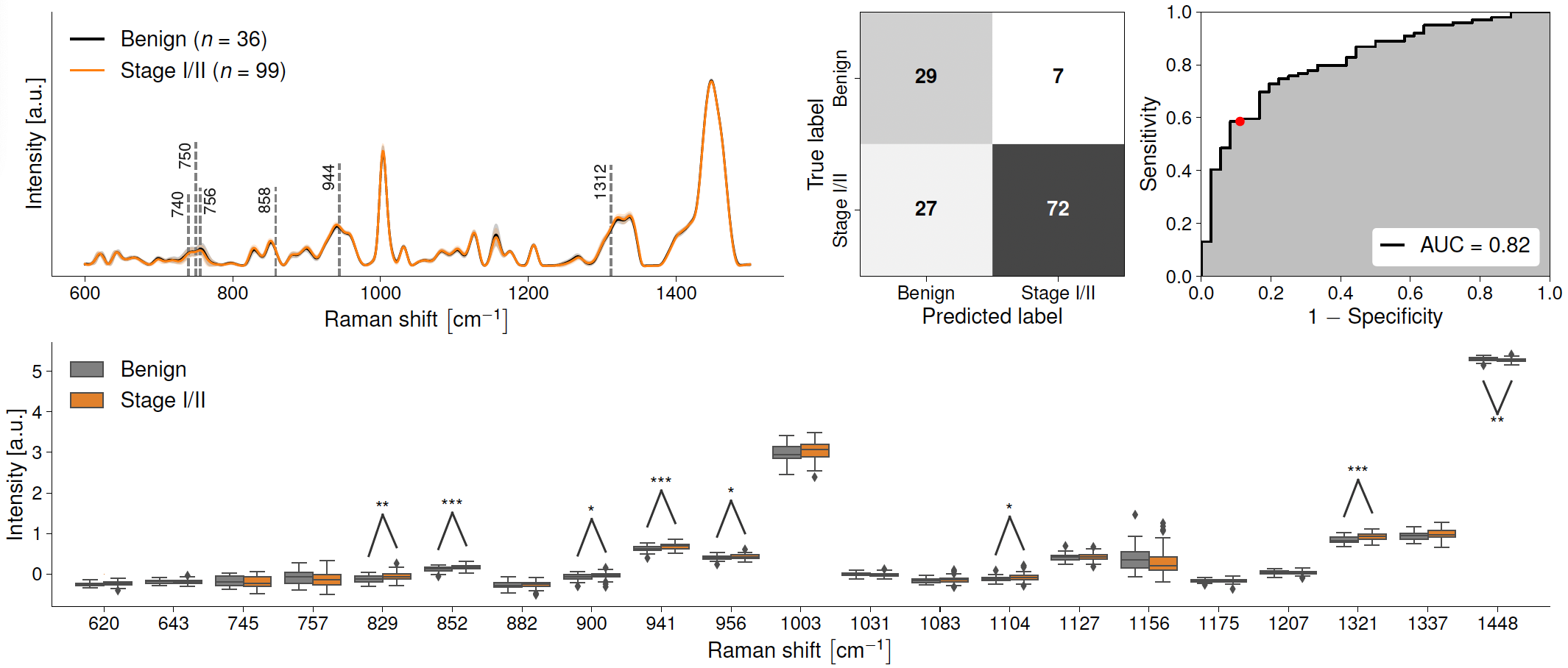

**Supplementary Figure 10: Raman spectroscopy detection of benign tumors vs. Stage I-II lung cancers (Model 13).** (A) Raman spectra of liquid blood plasma from patients with benign tumors (black line) and Stage I-II lung cancer (orange). Black dashed lines show peaks used in the machine learning models. (B) Confusion matrix showing efficacy of Raman spectral prediction of benign tumors and Stage I-II lung cancer in liquid blood plasma. (C) Receiver operating characteristic (ROC) curves for machine learning models discriminating between liquid blood plasma spectral fingerprints from the two groups. (D) Box-and-whisker plot showing the relative intensities of different Raman bands between liquid blood plasma from patients with benign tumors (grey) and Stage I-II lung cancer (orange). * represents a p-value of ≤0.05. ** represents a p-value of ≤ 0.01. *** represents a p-value of ≤ 0.001.

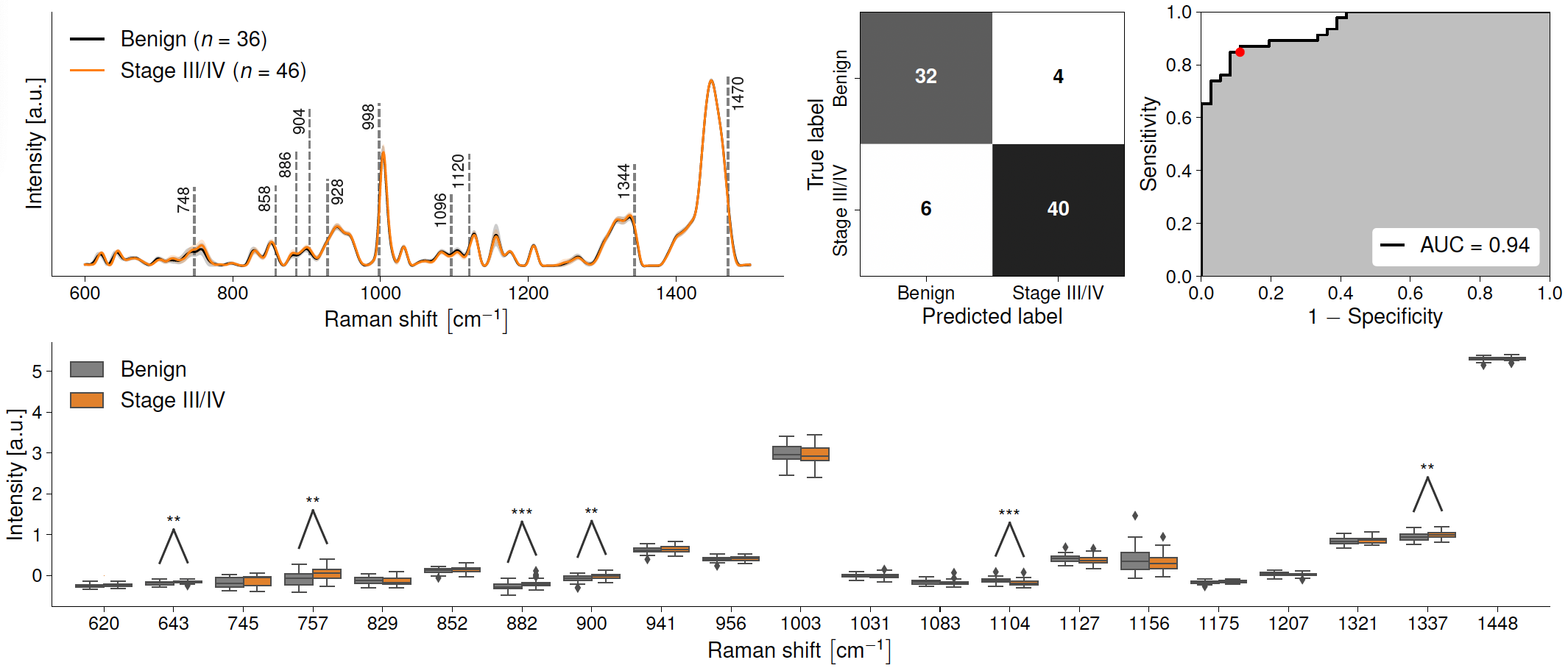

**Supplementary Figure 11: Raman spectroscopy detection of benign tumors vs. Stage III-IV lung cancers (Model 14).** (A) Raman spectra of liquid blood plasma from patients with benign tumors (black line) and Stage III-IV lung cancer (orange). Black dashed lines show peaks used in the machine learning models. (B) Confusion matrix showing efficacy of Raman spectral prediction of benign tumors and Stage III-IV lung cancer in liquid blood plasma. (C) Receiver operating characteristic (ROC) curves for machine learning models discriminating between liquid blood plasma spectral fingerprints from the two groups. (D) Box-and-whisker plot showing the relative intensities of different Raman bands between liquid blood plasma from patients with benign tumors (grey) and Stage III-IV lung cancer (orange). * represents a p-value of ≤0.05. ** represents a p-value of ≤ 0.01. *** represents a p-value of ≤ 0.001.

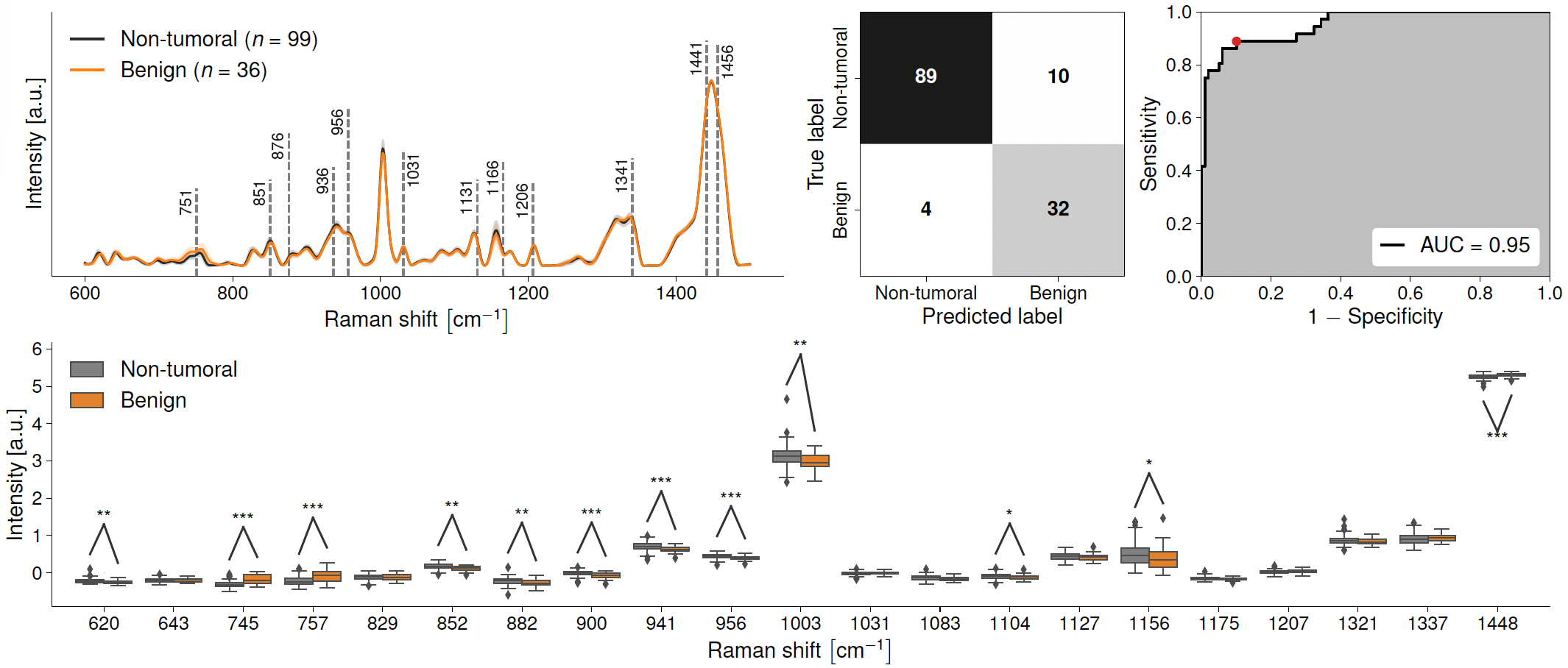

**Supplementary Figure 12: Raman spectroscopy detection of non-tumoral lung pathologies vs. benign tumors (Model 15).** (A) Raman spectra of liquid blood plasma from non-tumoral lung pathologies (black line) and benign tumors (orange). Black dashed lines show peaks used in the machine learning models. (B) Confusion matrix showing efficacy of Raman spectral prediction of non-tumoral lung pathologies and benign tumors in liquid blood plasma. (C) Receiver operating characteristic (ROC) curves for machine learning models discriminating between liquid blood plasma spectral fingerprints from the two groups. (D) Box-and-whisker plot showing the relative intensities of different Raman bands between liquid blood plasma from non-tumoral lung pathologies (grey) and those with benign tumors (orange). * represents a p-value of ≤0.05. ** represents a p-value of ≤ 0.01. *** represents a p-value of ≤ 0.001.

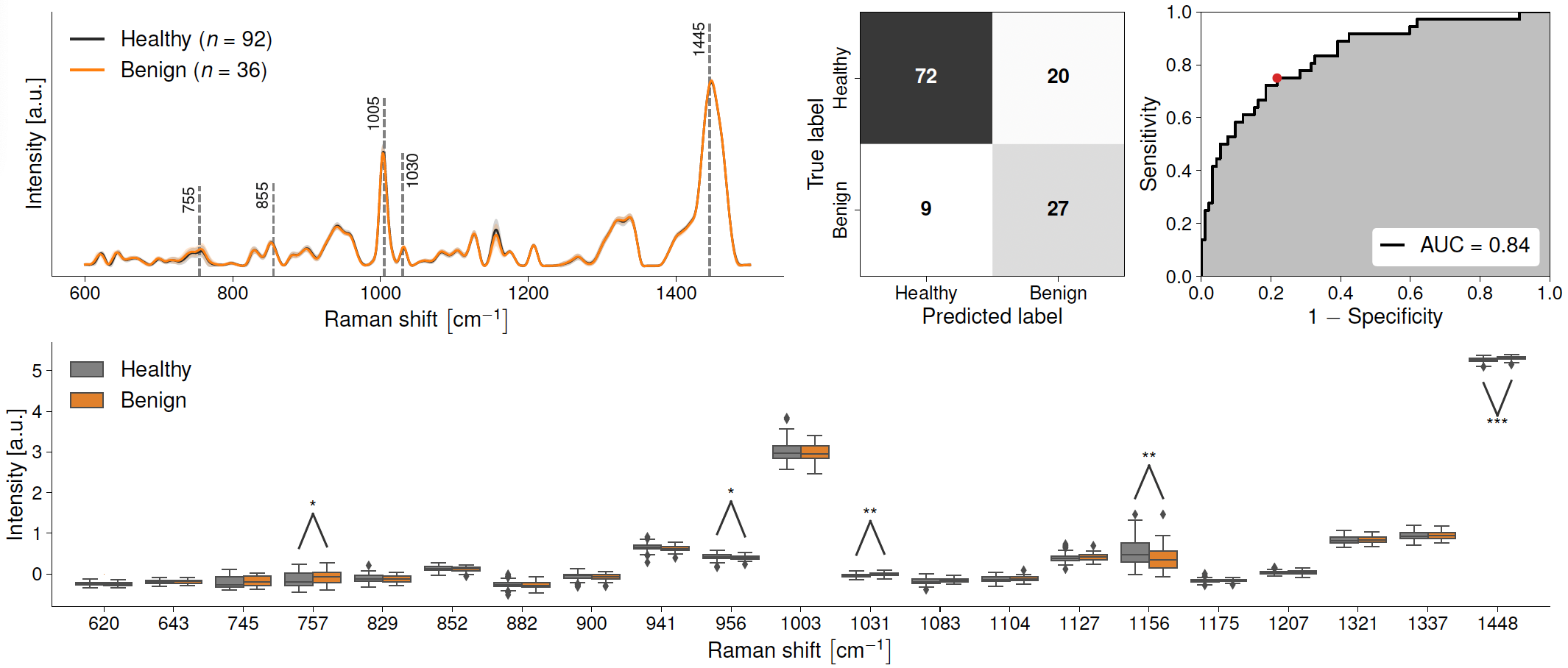

**Supplementary Figure 13: Raman spectroscopy detection of healthy controls vs. benign tumors (Model 16).** (A) Raman spectra of liquid blood plasma from healthy controls (black line) and benign tumors (orange). Black dashed lines show peaks used in the machine learning models. (B) Confusion matrix showing efficacy of Raman spectral prediction of healthy controls and benign tumors in liquid blood plasma. (C) Receiver operating characteristic (ROC) curves for machine learning models discriminating between liquid blood plasma spectral fingerprints from the two groups. (D) Box-and-whisker plot showing the relative intensities of different Raman bands between liquid blood plasma from healthy controls (grey) and those with benign tumors (orange). * represents a p-value of ≤0.05. ** represents a p-value of ≤ 0.01. *** represents a p-value of ≤ 0.001.

**Smoking Status Assessment**

Machine learning models were built to discriminate between different smoking status groups within the same pathology group. For example, a machine learning model was built to classify non-smokers and smokers in the non-tumoral group as this had enough spectra in each class (**Model 17,** **Supplementary Figure 14**). The AUC was 0.78, accuracy 74%, sensitivity 74% and specificity 74%. The only two Raman peaks that were statistically different in intensity were 643 (protein, tyrosine) which increased and 1156 cm^-1^ (carotenoids) which decreased. These two peaks were also present in models 7-10, 14 and 16, and may contribute to the “lung cancer” Raman spectrum in stages III-IV lung cancer. However, the peak at 643 cm^-1^ was not present in Model 4, which differentiates between control and stage I-II lung cancer. It has been established that smokers have statistically lower serum beta-carotene. This may be due to dietary factors or chemical breakdown of carotenoids by cigarette smoke.^64–66^

**Model 18** (**Supplementary Figure 15)** classified non-smokers and ex-smokers within the healthy control cohort. The AUC was low, at 0.66, accuracy 69%, sensitivity 75% and specificity 61%. The only Raman peaks that were statistically different in intensity were 1321 and 1337 cm^-1^ - both in the same protein band associated with C-H bonds, and both of which decreased. This trend was not seen in any of the machine learning models 1-16 in the positive group compared to the negative group, suggesting that being an ex-smoker is not linked to the “lung cancer” Raman spectrum.

**Model 19** (**Supplementary Figure 16)** classified ex-smokers and smokers within the Stage I-II lung cancer cohort. The AUC was low, at 0.56, accuracy 58%, sensitivity 50% and specificity 67%. The only two Raman peaks that were statistically different in intensity were 620 (protein, amino acids) and 882 cm^-1^, both of which decreased.

Model 20 (**Supplementary Figure 17**) incorporated only ex-smokers and discriminated between healthy controls and Stage III-IV ex-smokers. This was the equivalent model to Model 5, but with only data from ex-smokers. The key features that were used by machine learning model 20 were the identical to those used by machine learning model 5. For Model 5, the AUC was 0.95, accuracy was 88%, sensitivity 87% and specificity 89%. For Model 20, the AUC was 0.9, accuracy was 81%, sensitivity was 79% and specificity 83%. This slight drop in accuracy may be due to confounding variable of smoking or due to the 37% reduction in available data (n=138 vs n=86).

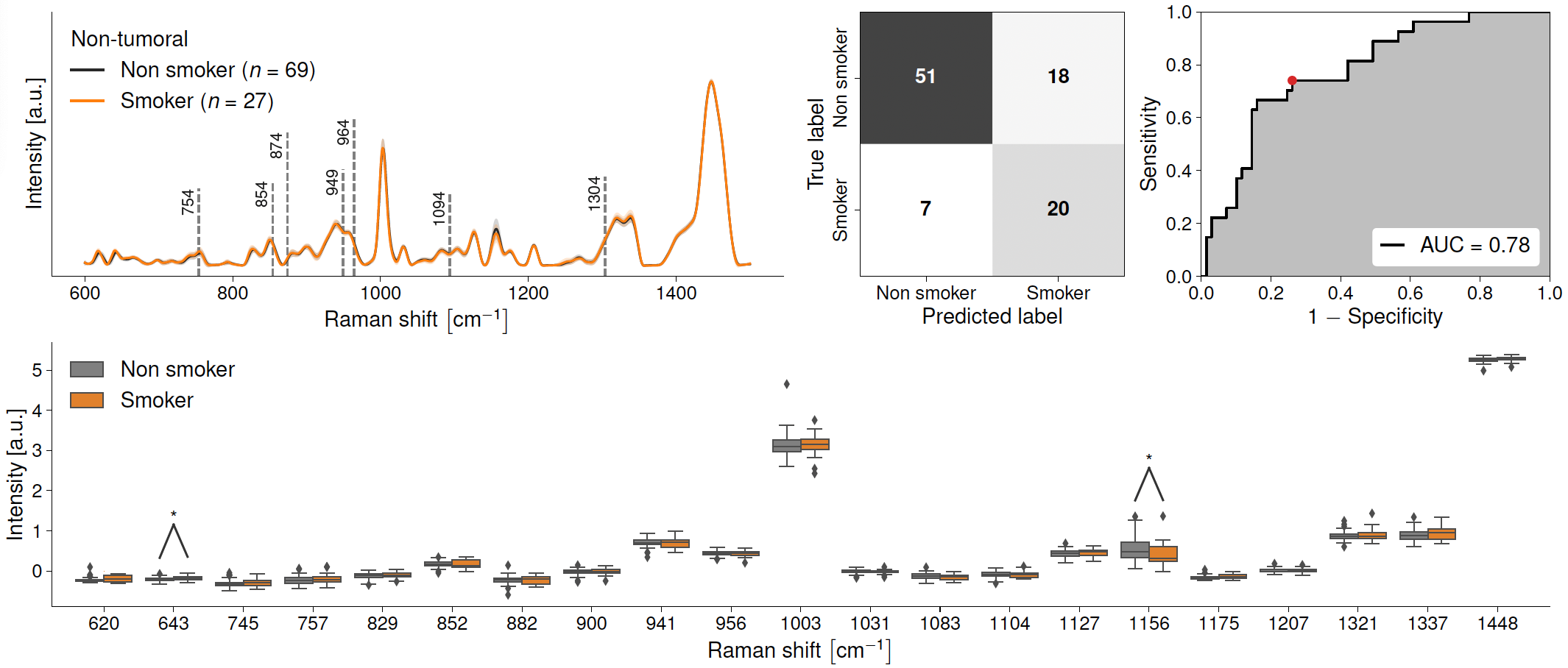

**Supplementary Figure 14: Raman spectroscopy detection of non-tumoral non-smokers vs. non-tumoral smokers (Model 17).** (A) Raman spectra of liquid blood plasma from non-tumoral non-smokers (black line) and non-tumoral smokers (orange). Black dashed lines show peaks used in the machine learning models. (B) Confusion matrix showing efficacy of Raman spectral prediction of non-tumoral non-smokers and non-tumoral smokers in liquid blood plasma. (C) Receiver operating characteristic (ROC) curves for machine learning models discriminating between liquid blood plasma spectral fingerprints from the two groups. (D) Box-and-whisker plot showing the relative intensities of different Raman bands between liquid blood plasma from non-tumoral non-smokers (grey) and those with non-tumoral smokers (orange). * represents a p-value of ≤0.05. ** represents a p-value of ≤ 0.01. *** represents a p-value of ≤ 0.001.

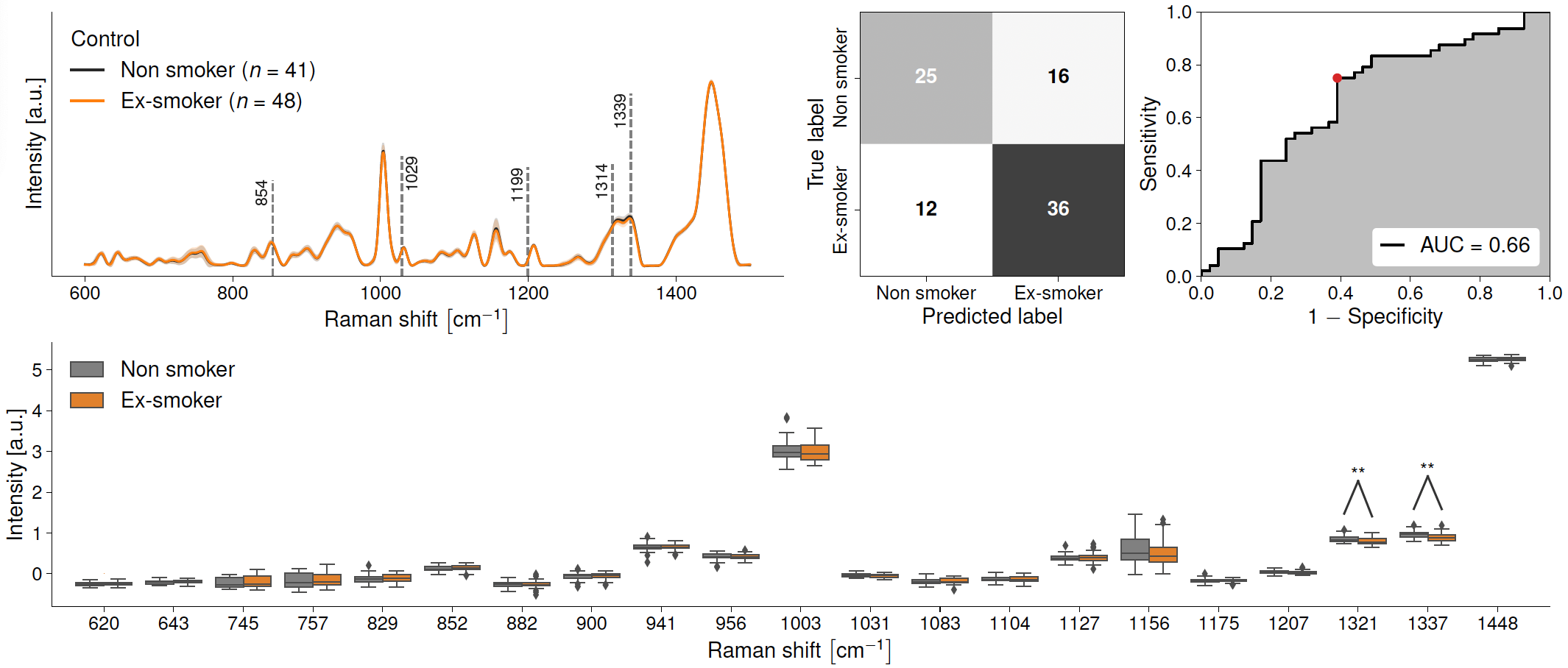

**Supplementary Figure 15: Raman spectroscopy detection of control non-smokers vs. control ex-smokers (Model 18).** (A) Raman spectra of liquid blood plasma from control non-smokers (black line) and control ex-smokers (orange). Black dashed lines show peaks used in the machine learning models. (B) Confusion matrix showing efficacy of Raman spectral prediction of control non-smokers and control ex-smokers in liquid blood plasma. (C) Receiver operating characteristic (ROC) curves for machine learning models discriminating between liquid blood plasma spectral fingerprints from the two groups. (D) Box-and-whisker plot showing the relative intensities of different Raman bands between liquid blood plasma from control non-smokers (grey) and those with control ex-smokers (orange). * represents a p-value of ≤0.05. ** represents a p-value of ≤ 0.01. *** represents a p-value of ≤ 0.001.

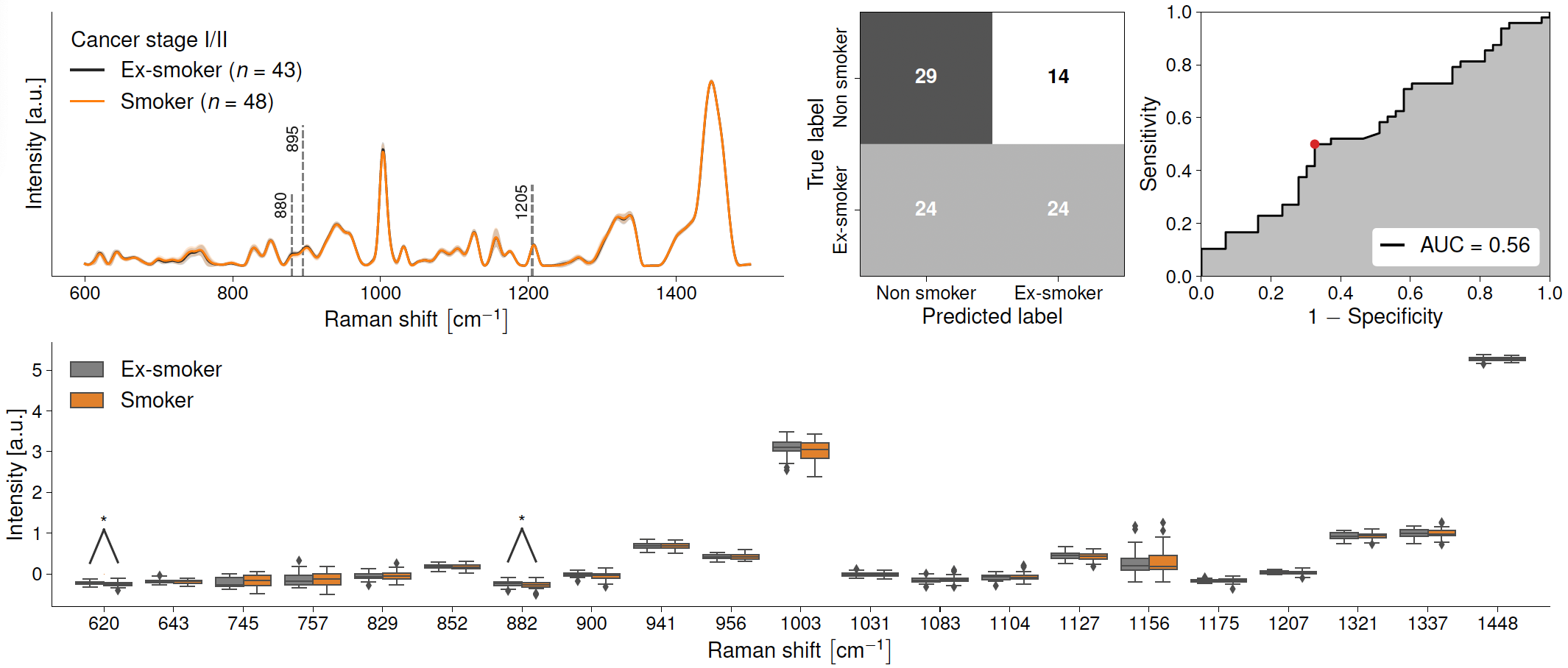

**Supplementary Figure 16: Raman spectroscopy detection of cancer stage I/II ex-smokers vs. cancer stage I/II smokers (Model 19).** (A) Raman spectra of liquid blood plasma from cancer stage I/II ex-smokers (black line) and cancer stage I/II smokers (orange). Black dashed lines show peaks used in the machine learning models. (B) Confusion matrix showing efficacy of Raman spectral prediction of cancer stage I/II ex-smokers and cancer stage I/II smokers in liquid blood plasma. (C) Receiver operating characteristic (ROC) curves for machine learning models discriminating between liquid blood plasma spectral fingerprints from the two groups. (D) Box-and-whisker plot showing the relative intensities of different Raman bands between liquid blood plasma from cancer stage I/II ex-smokers (grey) and those with cancer stage I/II smokers (orange). * represents a p-value of ≤0.05. ** represents a p-value of ≤ 0.01. *** represents a p-value of ≤ 0.001.

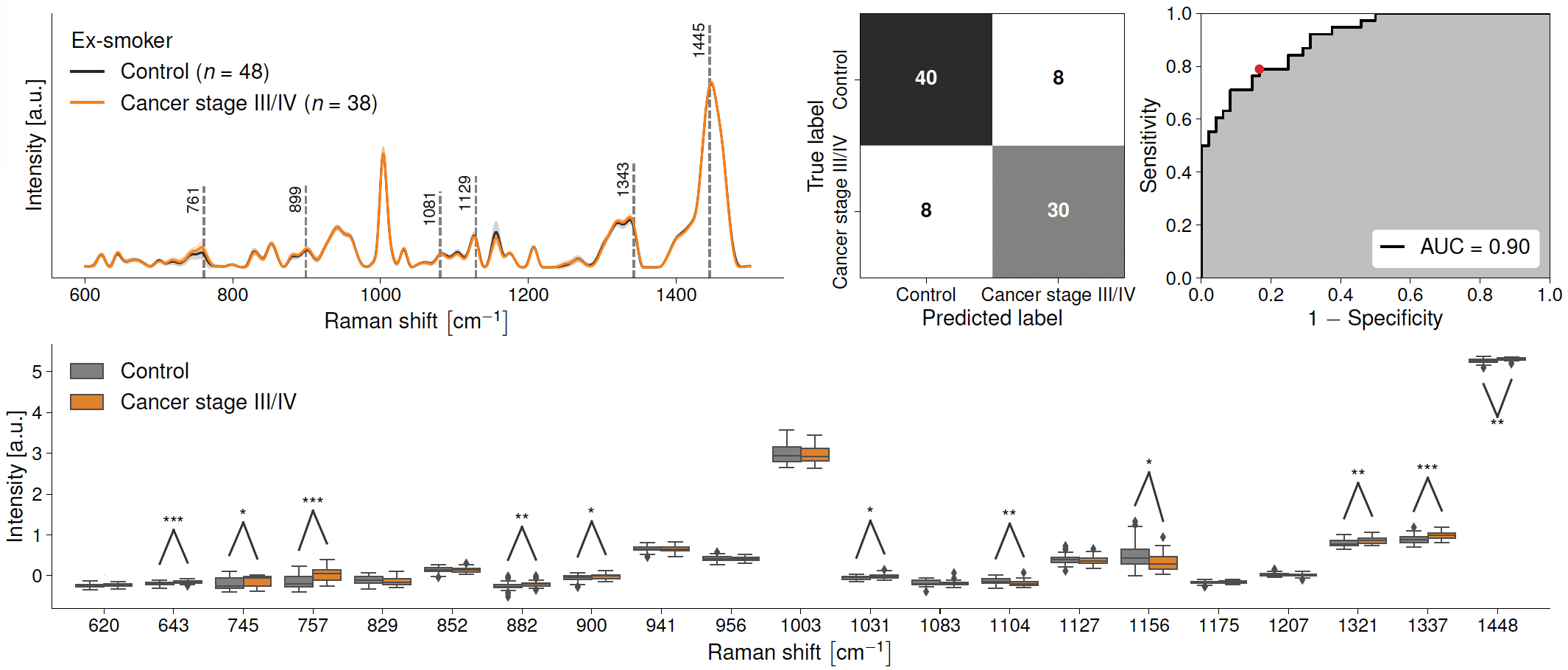

**Supplementary Figure 17: Raman spectroscopy detection of ex-smoker healthy controls vs. cancer stage I/II smokers (Model 20).** (A) Raman spectra of liquid blood plasma from cancer stage I/II ex-smokers (black line) and cancer stage I/II smokers (orange). Black dashed lines show peaks used in the machine learning models. (B) Confusion matrix showing efficacy of Raman spectral prediction of cancer stage I/II ex-smokers and cancer stage I/II smokers in liquid blood plasma. (C) Receiver operating characteristic (ROC) curves for machine learning models discriminating between liquid blood plasma spectral fingerprints from the two groups. (D) Box-and-whisker plot showing the relative intensities of different Raman bands between liquid blood plasma from cancer stage I/II ex-smokers (grey) and those with cancer stage I/II smokers (orange). * represents a p-value of ≤0.05. ** represents a p-value of ≤ 0.01. *** represents a p-value of ≤ 0.001.
